# Effective connectivity extracts clinically relevant prognostic information from resting state activity in stroke

**DOI:** 10.1101/2020.12.11.20247783

**Authors:** Mohit H Adhikari, Joseph Griffis, Joshua S. Siegel, Michel Thiebaut de Schotten, Gustavo Deco, Andrea Instabato, Mathieu Gilson, Maurizio Corbetta

**Author notes:** Equal contribution (Mathieu Gilson, Maurizio Corbetta). Corresponding author: Mohit H Adhikari, Center for Brain and Cognition, University of Pompeu Fabra, Calle Ramon Trias Fargas 25-27, Barcelona 08005.

## Abstract

Recent resting-state fMRI studies in stroke patients have identified two robust biomarkers of acute brain dysfunction: a reduction of inter-hemispheric functional connectivity (FC) between homotopic regions of the same network, and an abnormal increase of ipsilesional FC between task-negative and task-positive resting-state networks (RSNs). Whole-brain computational modeling studies, at the individual subject level, using undirected effective connectivity (EC) derived from empirically measured FC, have shown a reduction of measures of integration and segregation in stroke as compared to healthy brains. Here we employ a novel method, first, to infer whole-brain directional EC from zero-lagged and lagged FC, then, to compare it to empirically measured FC for predicting stroke vs. healthy status, and patient performance (zero, one, multiple deficits) across neuropsychological tests. We also investigated the accuracy of FC vs. model EC in predicting the long-term outcome from acute measures.

Both FC and EC predicted healthy from stroke individuals significantly better than the chance-level, however, EC accuracy was significantly higher than that of FC at 1-2 weeks, three months, and one year post-stroke. The predictive FC links mainly included those reported in previous studies (within-network inter-hemispheric, and between task-positive and -negative networks intra-hemispherically). Predictive EC links included additional between-network links. EC was a better predictor than FC of the number of behavioral domains in which patients suffered deficits, both at two weeks and one-year post onset of stroke. Interestingly, patient deficits at one-year time point were better predicted by EC values at two weeks rather than at one-year time point. Our results thus demonstrate that the second-order statistics of fMRI resting-state activity at an early stage of stroke, derived from a whole-brain EC, estimated in a model fitted to reproduce the propagation of BOLD activity, has pertinent information for clinical prognosis.

## INTRODUCTION

First-time stroke patients with heterogeneous lesion location show widespread alterations of resting-state functional connectivity (FC) − the temporal correlation of the fMRI signals across brain regions and networks - in the acute stage (He *et al*., 2007; Carter *et al*., 2010, 2012; Baldassarre *et al*., 2014), what is now called ‘connectional diaschisis’ (Carrera and Tononi, 2014). The most common abnormalities include a reduction of the inter-hemispheric FC between homotopic regions of the same networks, and an increase of intra-hemispheric FC between regions/networks that are typically not or negatively correlated (e.g., task-positive and task-negative networks)(Siegel *et al*., 2016). Abnormalities of FC are strongly correlated with acute behavioral deficits and their recovery(Siegel *et al*., 2016, 2018). The low dimensionality of FC alterations across brain regions has been proposed as as an explanation for why behavioral deficits are also low dimensional and correlated across subjects, with three sets of deficit components accounting for ∼70% of the behavioral variance(Corbetta *et al*., 2015, 2018).

We hypothesize that stroke lesions induce an abnormal state of low neural and behavioral variability. Accordingly, whole-brain network models of sub-acute stroke have found that decrement of integration and segregation measured as the entropy of potential spatial FC topography correlated with these two FC based markers of stroke, and with behavioral deficits(Adhikari *et al*., 2017). Adhikari et al. used in their whole-brain model a group average structural connectivity optimized in each subject (healthy, stroke) by inferring the symmetrical effective connectivity (EC) from each patient’s own FC using a heuristic, gradient descent approach. The effect of lesions was modeled by weakening the structural edges to/from a damaged cortical region.

A weakness of this approach is that FC captures the levels of correlated activity between a pair of brain regions, but it does not describe the causal influence from one region to another or the simultaneous influence of a third region. A second weakness is that stroke lesions affect not only cortical parcels but also predominantly white matter pathways (Corbetta et al. 2015). Therefore, a better approach for estimating the effect of lesions would be to take in account the structural disconnection, between both damaged and non-damaged regions, caused by lesions.

In contrast to FC, effective connectivity (EC) measures directed interactions and indirectly, some form of causality. Several methods, such as Granger causality (Goebel *et al*., 2003), dynamic causal modeling (Friston *et al*., 2014), partial correlations (Smith *et al*., 2011) have been developed to infer effective connectivity. Recently, Gilson et al. (Gilson *et al*., 2016) have demonstrated a theoretically robust technique to infer whole-brain EC from zero-lagged and lagged resting-state FC time-series. Modeling each local brain region with an Ornstein-Uhlenbeck process, the inference procedure uses a mask of the putatively existing anatomical links as a topological prior and since it also estimates non-symmetric lagged FC, the resulting EC is directional as well as sparse (reflecting anatomically plausible links). EC changes were found to align with those known from electrophysiology in the visual field mapping between early visual cortices (Gravel *et al*., 2020). Pallarés *et al*., (2018) showed that EC predicts in a cross-validated fashion the identity of individual healthy participants from fMRI sessions more accurately than FC.

In this study, we first obtained the EC for healthy participants and first-time stroke patients, using the approach of Gilson et al., at three time points post onset of stroke: two weeks, three months and one year. Critically, the disconnection effect of lesions was estimated embedding the volume of each lesion in an atlas of normal white matter connections (Yeh *et al*., 2018), and estimating the severity of disconnection among cortical regions. Then, following the methodology of Pallares et al., we used a multinomial logistic regression (MLR) classifier to classify this cohort in multiple classification using EC or FC values for all links present in the atlas of Yeh et al. The first classification diagnosed patients from healthy controls at each time point using same time point EC and FC values of links (e.g., two weeks EC or FC values of links to distinguish two weeks stroke patients from healthy participants).

The second classification aimed to diagnose stroke individuals with different numbers of behavioral deficits (zero, one, many) across seven domains of impairments using EC and FC values of links at the same time-point. Besides, we tried EC and FC of links from previous time points to predict future behavioral deficit status (e.g., use two weeks FC or EC values to predict three months and one year time point behavioral scores).

We hypothesized that EC would be a more sensitive biomarker than FC, especially for more dimensional behavioral predictions. This hypothesis is based on EC’s theoretical ability to infer directional interactions and its superiority (as compared to FC) in predicting individual scans (Pallarés *et al*., 2018). Moreover, we hypothesized that EC predictive topography will provide additional and complementary information to FC topography.

## MATERIALS AND METHODS

### Stroke sample

Subjects (n=172) were prospectively recruited, of whom 132 met post-enrollment inclusion criteria (for details see Corbetta *et al*., 2015).

#### Inclusion criteria

1. Age 18 or greater. No upper age limit. 2. First symptomatic stroke, ischemic or hemorrhagic. 3. Up to two lacunes, clinically silent, less than 15 mm in size on CT scan. 4. Clinical evidence of motor, language, attention, visual, or memory deficits based on neurological examination. 5. Time of enrollment: <2 weeks from stroke onset. 6. Awake, alert, and capable of participating in research.

#### Exclusion criteria

1. Previous stroke based on clinical imaging. 2. Multi-focal strokes. 3. Inability to maintain wakefulness in the course of testing. 4. Presence of other neurological, psychiatric or medical conditions that preclude active participation in research and/or may alter the interpretation of the behavioural/imaging studies (e.g. dementia, schizophrenia), or limit life expectancy to less than 1 year (e.g. cancer or congestive heart failure class IV). 5. Report of claustrophobia or metal object in body.

### Healthy controls

A healthy control group (N=25) were matched with the study sample for age, gender, and years of education.

### Lesion identification

Lesions were manually segmented on each patient’s structural MRI scans using the Analyze software package (Robb and Hanson, 1991). Surrounding vasogenic edema was included in the lesion definition for patients with hemorrhagic stroke. All segmentations were reviewed by two board certified neurologists (Maurizio Corbetta and Alexandre Carter), and were reviewed a second time by MC for scoring of number of lacunes and periventricular white matter disease. The final segmentations were used as binary lesion masks for subsequent processing and analysis steps. Lesion masks were transformed to MNI atlas space using a combination of linear transformations and non-linear warps and were resampled to have isotropic voxel resolution.

### Behavioral testing

All subjects and controls underwent a behavioral battery that included assessment of motor, language, attention, memory, and visual function following each scanning session. Overall clinical deficit was also assessed in each patient using the NIH stroke scale (NIHSS) (Brott *et al*., 1989). Imaging and behavioral testing session usually were performed on the same day. Dimensionality reduction was performed on the behavioral performance data as described previously (Corbetta *et al*., 2015). Principal components analysis was performed on all tests within a behavioral domain to produce a single score that predicted the majority of variance across tasks. The ‘Motor’ score describes contralesional deficits that correlated across shoulder flexion, wrist extension/flexion, ankle flexion, hand dynamometer, nine hole peg, action research arm test, timed walk, functional independence measure, and the lower extremity motricity index. The ‘Visual Field Attention’ score describes contra-lesional attention biases in Posner, Mesulam, and BIT center of cancellation tasks. A separate ‘Sustained Attention’ score loaded on non-spatial measures of overall performance, reaction time, and accuracy on the same tests. A third ‘Shifting Attention’ score loaded on tests indexing attention shifts, e.g. the difference in response times for attended vs. unattended targets. The ‘Spatial Memory’ score loaded on the Brief Visuospatial Memory Test and spatial span. The ‘Verbal Memory’ score loaded on the Hopkins Verbal Learning Test. The ‘Language’ score loaded on both comprehension (complex ideational material, commands, reading comprehension) and production (Boston naming, oral reading).

We normalized the score of each of the seven factors for each patient using the mean and standard deviation of the corresponding factor scores in age-matched healthy controls. KNN imputation was employed to impute the scores for the missing domains in a patient using weighted average across 5 of its nearest-neighbours. A z-score of −2 or less, that is, a factor score of at least two standard deviation below average score in healthy controls, was considered to be a deficit.

### fMRI procedure and scanning

Patients were scanned at three time points – two weeks (mean = 13.4 days, SD = 4.8 days), three months (mean = 112.5 days, SD = 18.4 days) and one-year (mean = 393.5 days, SD = 55.1 days) post onset of stroke. Healthy controls were scanned at two time points separated by 3 month interval.

Scanning was performed with a Siemens 3T Tim-Trio scanner at the School of Medicine of the Washington University in St. Louis. All participants underwent structural, functional and diffusion tensor scans. Structural scans consisted of: (1) a sagittal MP-RAGE T1-weighted image (TR=1950 msec, TE=2.26 msec, flip angle=90 deg, voxel size=1.0 x 1.0 x 1.0 mm, slice thickness = 1.00 mm); (2) a transverse turbo spin-echo T2-weighted image (TR=2500 msec, TE=435 msec, voxel-size=1.0 x 1.0 x 1.0 mm, slice thickness = 1.00 mm); and (3) a sagittal FLAIR (fluid attenuated inversion recovery) (TR=7500 msec, TE=326 msec, voxel-size=1.5 x 1.5 x 1.5 mm, Slice thickness = 1.50 mm). PASL acquisition parameters were: TR = 2,600 msec, TE = 13 msec, flip angle = 90°, bandwidth 2.232 kHz/Px, and FoV 220 mm; 120 volumes were acquired (322 s total), each containing 15 slices with slice thickness 6- and 23.7-mm gap. Resting state functional scans were acquired with a gradient echo EPI sequence (TR = 2,000 msec, TE = 27 msec, 32 contiguous 4-mm slices, 4 × 4 mm in-plane resolution) during which participants were instructed to fixate on a small cross in a low luminance environment. Six to eight resting state fMRI runs, each including 128 volumes (30 min total), were acquired.

### fMRI data preprocessing

Functional MRI data underwent a preprocessing procedure consisting of the following steps: 1) asynchronous slice acquisition was compensated by sinc interpolation to align all slices 2) elimination of odd/even slice intensity differences resulting from interleaved acquisition 3) a whole brain normalization corrected for changes in signal intensity across scans 4) data were realigned within and across scans to correct for head movement 5) EPI data were co-registered to the subject’s T2-weighted anatomical image, which in turn was co-registered with the T1-weighted MP-RAGE, in both cases using a cross-modal procedure based on alignment of image gradients (Rowland *et al*., 2005). The MP-RAGE was then transformed to an atlas-space (Talairach and Tournoux, 1988) representative target using a 12-parameter affine transformation. Movement correction and atlas transformation were accomplished in one resampling step (resulting in an isotropic 3 mm voxel size) to minimize blur and noise. In preparation for the FC MRI analysis, data were passed through several additional preprocessing steps (Fox *et al*., 2005): (1) spatial smoothing (6 mm full width at half maximum Gaussian blur), (2) temporal filtering retaining frequencies in the 0.009–0.08 Hz band, and (3) removal of the following sources of spurious variance unlikely to reflect spatially specific functional correlations through linear regression: (i) six parameters obtained by rigid body correction of head motion, (ii) the whole-brain signal averaged over a fixed region in atlas space, (iii) signal from a ventricular region of interest (ROI), and (iv) signal from a region centered in the white matter.

The decision to include whole-brain signal as a nuisance regressor was made in light of numerous studies showing that whole-brain signal regression removes substantial confound attributable to head motion and respiration (CO_2_) and reduces spurious correlations in BOLD data (Power *et al*., 2012, 2015; Satterthwaite *et al*., 2012; Van Dijk *et al*., 2012). A recently published report found that functional connectivity changes observed in stroke patients relative to age-matched controls were consistent with and without whole-brain signal regression (Siegel *et al*., 2016).

### Quality control of resting-state data

For each frame of resting state fMRI scans a DVARS (temporal derivative of timecourses of RMS variance over voxels) score was calculated. DVARS indexes the rate of change of the BOLD signal across the entire brain at each frame of data; the rationale and method for calculating DVARS has been described previously (Power *et al*., 2012). To define the DVAR threshold for our group of patients, we computed the mean plus 2 standard deviation of DVARS values for all frames, excluding the first 4 frames, in the group of age-matched control subjects. The threshold was equal to 4.6. All frames in the resting state fMRI scans with a DVARS value of 4.6 or higher were linearly interpolated over in order to maintain the temporal continuity for each scan.

Exclusion criteria for functional connectivity quality included 1) less than 180 usable frames, and 2) severe hemodynamic lags (greater than 500 msec inter-hemispheric difference) measured from RS-fMRI (Siegel *et al*., 2015). After motion and lag exclusion, 98 patients were included at two weeks, 74 patients at three months, 53 patients at one year, 23 controls at time point 1 and 22 control at time point 2.

### Parcels and system assignments

Functional connectivity and covariance matrices were obtained for all links between 324 cortical parcels from the Gordon333 cortical parcellation and resting-state network assignments (available at http://www.nil.wustl.edu/labs/petersen/Resources.html) This parcellation is derived from the RS-fMRI data from 120 healthy individuals (Gordon *et al*., 2016), and consists of 333 cortical parcels associated with 13 RSNs. Owing to very low number of vertices, 9 parcels were excluded as done in previous studies (Siegel *et al*., 2016, 2018; Griffis *et al*., 2019). The remaining 324 surface-based cortical parcels were assigned to these 13 RSNs: visual (VIS), retro-splenial (RSP), somato-motor hand (SMH), somato-motor mouth (SMM), auditory (AUD), cingulo-opercular (COP), ventral attention network (VAN), salience (SAL), cingulo-parietal (CPL), dorsal attention network (DAN), frontal parietal network (FPN), default mode network (DMN) and none of the above (NON).

### Mask of neuroanatomical structural links for EC estimation

As mentioned in the introduction, the effective connectivity is estimated for a mask of strongest neuroanatomical links for each individual participant. This mask referred to as EC_mask_Ctl was identical for each healthy control participant and was formed by the union of two sets. The first set included all links stronger than 0.5% of the strongest link in the structural connectivity matrix (SC-Ctl) averaged across those of healthy control participants of this study. Individual healthy control SC matrices were obtained using diffusion weighted imaging (DWI) and probabilistic tractography as described in the next section. The second set included links present in the structural connectome template created from a publicly available tractography atlas (Yeh *et al*., 2018), constructed using DWI data from 842 Human connectome project (HCP) participants and deterministic tractography. We henceforth refer to this structural connectome template as HCP-SCT. These two sets of links were not disjoint and some links were common. The mask of neuroanatomical links considered for EC estimation was individualized in each patient depending on the impact of lesion. Thus EC_mask_Pat_i_, for the ith patient, included all links in EC_mask_Ctl except, those from and to a ROI with 100% grey matter damage and all other completely damaged links in the patient’s structural disconnectome (SDC).

The rationale for including in EC_mask_Ctl both the strongest links in SC-Ctl as well as those present in HCP-SCT was many-fold. First, due to the end-to-end nature of tractography used by Yeh et al., the links in the HCP-SCT were very reliable but too few (N=4218), vis-à-vis the high spatial resolution of the parcellation, to obtain a robust estimate of EC that can produce sufficiently high fit between model and data (Gilson *et al*., 2016, 2019). Therefore it was necessary to include the strongest links in SC-Ctl (N=28256) in the estimation of EC. However, while they were sparse, links present in HCP-SCT were found to have stronger FC, on average, in both controls and patients than those not present in HCP-SCT(Griffis *et al*., 2020). Thus the sparseness and reliability of links in the HCP-SCT allowed us to use only them for classification and biomarker identification, as will be explained in the subsection ‘Classification’. A practical reason for this choice is to keep the computational cost low without losing the prediction accuracy (Figure S1). A more clinically oriented reason is to seek the effect of strokes among the most stable part of the structural backbone of the brain connectivity, independently of individual variability in SC as well as patient-specific stroke characteristics. Also, incorporating impact of lesions on white matter pathways through structural disconnections for each individual patient was possible only through the HCP-SCT.

Methods for generation of SC-Ctl, HCP-SCT as well as SDC for individual patients are described in the following sections.

### Structural connectivity for healthy controls (SC-Ctl)

#### Acquisition

A total of 64 near-axial slices were acquired on a Siemens Trio scanner equipped with a 12ch head coil. We used a fully optimized acquisition sequence for the tractography of DWI, which provided isotropic (2 × 2 × 2 mm) resolution and coverage of the whole head with a posterior-anterior time point of acquisition. We used a repetition time (TR) equivalent to 9.2s. At each slice location, 4 images were acquired with no diffusion gradient applied. Additionally, 60 diffusion-weighted images were acquired, in which gradient directions were uniformly distributed on the hemisphere with electrostatic repulsion. The diffusion weighting was equal to a b-value of 1000 sec mm−2. In order to optimize the contrast of acquisition, this sequence was repeated twice.

#### Processing

For each slice, diffusion-weighted data were simultaneously registered and corrected for subject motion and geometrical distortion adjusting the diffusion directions accordingly (ExploreDTI http://www.exploredti.com; Leemans and Jones, 2009)

Spherical deconvolution was chosen to estimate multiple orientations in voxels containing different populations of crossing fibres (Tournier *et al*., 2004; Alexander, 2005; Anderson, 2005). The damped version of the Richardson-Lucy algorithm for spherical deconvolution (Dell’acqua *et al*., 2010) was calculated using Startrack (https://www.mr-startrack.com)

Algorithm parameters were chosen as previously described (Dell’Acqua *et al*., 2013). A fixed fibre response corresponding to a shape factor of α = 1.5 × 10– 3 mm2/s was chosen (Dell’Acqua *et al*., 2013). Fiber orientation estimates were obtained by selecting the orientation corresponding to the peaks (local maxima) of the fibre orientation distribution (FOD) profiles. To exclude spurious local maxima, we applied both an absolute and a relative threshold on the FOD amplitude. A first “absolute” threshold was used to exclude intrinsically small local maxima due to noise or isotropic tissue. This threshold was set to 3 times the mean amplitude of a spherical FOD obtained from a grey matter isotropic voxel (and therefore also higher than an isotropic voxel in the cerebrospinal fluid). A second “relative” threshold of 10% of the maximum amplitude of the FOD was applied to remove the remaining local maxima with values higher than the absolute threshold (Dell’acqua *et al*., 2010).

Whole-brain tractography was performed selecting every brain voxel with at least one fibre orientation as a seed voxel. From these voxels, and for each fibre orientation, streamlines were propagated using Euler integration with a step size of 1 mm (as described in Dell’Acqua *et al*., 2013). When entering a region with crossing white matter bundles, the algorithm followed the orientation vector of least curvature (as described in Schmahmann *et al*., 2007) Streamlines were halted when a voxel without fibre orientation was reached or when the curvature between two steps exceeded a threshold of 60°.

#### Normalization

The whole-brain streamlines were registered to the standard MNI. For each participant, whole-brain streamline tractography was converted into streamline density volumes where the intensities corresponded to the number of streamlines crossing each voxel. A study-specific template of streamline density volumes was generated using the Greedy symmetric diffeomorphic normalization (GreedySyN) pipeline distributed with Advanced Normalization Tools (ANTs, http://stnava.github.io/ANTs/; Avants et al. 2011). This provided an average template of the streamline density volumes for all participants. The template was then co-registered with a standard 2 mm MNI152 template using flirt tool implemented in FSL to obtain a streamline density template in the MNI152 space. Finally, individual streamline density volumes were registered to the streamline density template in the MNI152 space template using ANTs GreedySyn. The same registration parameters were applied to the individual whole-brain streamline tractography using the trackmath tool distributed with the software package Tract Querier (Wassermann *et al*., 2016) using ANTs GreedySyn. This step produced a whole-brain streamline tractography in the standard MNI152 space.

#### Dissections

Dissections were performed using trackvis (http://trackvis.org; Wedeen *et al*., 2008). Regions of interest were derived from Gordon et al. (2016) and arranged 2 by 2 in order to select streamlines and build a connectivity matrix for each participant. We took the number of streamlines existing between two ROIs divided by total number of vertices in two ROIs as a measure of the strength of the structural connection between them.

The SC matrices constructed in this manner for all healthy participants were averaged to construct an average SC matrix for healthy controls (SC-Ctl).

### Human Connectome Project derived structural connectome template (HCP-SCT)

We constructed the structural connectome template using the HCP-842 streamline tractography atlas (Yeh *et al*., 2018) as described in detail in a previous publication (Griffis *et al*., 2019). In brief, Yeh et al. (2018) performed deterministic fiber tracking (Yeh *et al*., 2013) on the high-angular resolution diffusion MRI data from 842 Human Connectome Project participants and extracted 550,000 streamline trajectories in MNI space that were then vetted and labelled by a team of neuroanatomists, and associated with 1 of 66 neuroanatomically defined fiber bundles (e.g., superior longitudinal fasciculus, corpus callosum, etc.) corresponding to commissural, association, projection, brainstem, and cerebellar pathways (cranial nerves were not included). As described in (Griffis *et al*., 2019), the copus callosum was split into 5 segments resulting in a total of 70 tracts. A 324×324 structural connectivity adjacency matrix A^S^ was then constructed where each entry A^S^_ij_ indexed the number of streamlines connecting region i and region j. Due to the close proximity of ventral visual and dorsal cerebellar regions, a small number of dorsal cerebellar streamlines were captured by the dilated visual regions. Therefore, we removed any links between visual areas and the cerebellum.

### Structural lesion features & structural disconnectome (SDC)

As described in detail in Griffis *et al*., 2019, parcel-wise grey matter damage measure was obtained for each patient by computing the proportion of each grey matter parcel that overlapped with each patient’s lesion. Subsequently, the expected disconnection for each patient was obtained by intersecting their MNI-registered lesion and the HCP-SCT. Therefore, for each patient, all streamlines from the 324×324 SC adjacency matrix A^S^, that passed through the lesion were extracted to obtain a 324×324 structural disconnection adjacency matrix *A*^*D*^ where each entry *A*^*D*^_*ij*_ indexed the number of streamlines connecting parcel *i* and parcel *j* that intersected the lesion (i.e. that were disconnected in that patient). Each element in matrix *A*^*D*^ was normalized by the corresponding value in *A*^*S*^, to obtain the proportion of streamlines connecting every pair of parcels that were disconnected by the lesion. This step accounted for differences in the number of streamlines connecting different parcel pairs and ensured that all disconnection measurements were directly comparable and intuitively interpretable in terms of proportional disconnection rather than number of streamlines (Griffis *et al*., 2019).

### Estimation of effective connectivity (EC)

We obtained whole-brain effective connectivity using a recently developed method (Gilson *et al*., 2016, 2019). This method optimizes the effective weights of structural links between local brain regions (quantifying the causal interactions between them) using a multivariate Ornstein-Uhlenbeck process model for each region and zero-lag and lagged covariances obtained from the empirical data. Optimization is done using a gradient descent algorithm that minimises the model error between model and data covariances. The EC is estimated for a mask of strongest neuroanatomical links; other link weights are kept to zero. As mentioned before, the mask, EC_mask_Ctl, used for healthy participants included strongest links in the SC-Ctl and all existing links in the HCP-SCT. The mask, EC_mask_Pat_i,_ for an individual patient excluded all completely damaged links in that patient from the mask for healthy controls. The detailed procedure for the estimation of EC is as follows:

#### Estimation of lagged and zero-lagged covariances & FC

For every session of *T* time frames, we denote the BOLD time series by 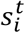 for each region 1 ≤ *i* ≤ *N* whose mean signal is denoted by 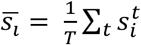. The empirical BOLD covariances to be optimized without and with time lag are then given by:

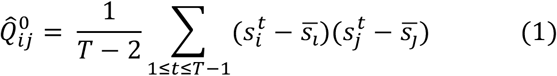

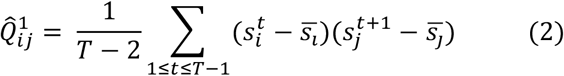

and the functional connectivity between region *i* and region *j* is given by:

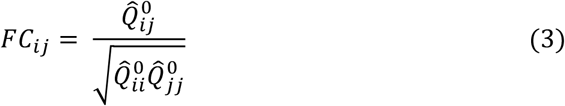

#### Multivariate Ornstein-Unlenbeck (MOU) process to model whole-brain dynamics

The activity variable, *x*_*i*_, for each region decays exponentially with a time constant, *τ*_*x*_ and evolves depending on the activity of other populations:

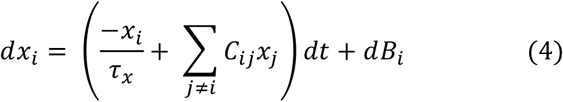

Here, the fluctuatung inputs, *dB*_*i*_, are independent and correspond to a diagonal covariance matrix Σ. In the model, all variables, *x*_*i*_, have zero mean. The spatiotemporal zero lagged and lagged covariances are denoted by 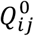 and 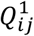 and can be calculated by solving the following consistency equations:

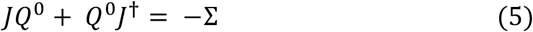

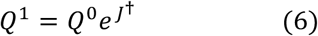

Here *J* is the Jacobian of the dynamical system; 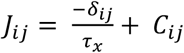, where, *δ*_*ij*_ is the Kronecker delta function and † denotes matrix transpose.

#### Lyapunov optimization for parameter estimation

Aim of the optimization procedure is to tune model parameters *C*_*ij*_ so that the model covariance matrices *Q*^0^ and *Q*^1^ reproduce the emprical ones 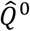 and 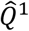. First, for every participant, we calculate the time constant *τ*_*ac*_ associated with the exponential decay of the autocovariance averaged across all ROIs using time lags between 0 and 2 TRs. The model is initialized with *τ*_*x*_= *τ*_*ac*_ and no connectivity (*C* = 0) as well as unit variances without covariances (Σ_*ij*_ = *δ*_*ij*_). That allows to calculate *J* and by solving equations (5) and (6), we get the model covariances *Q*^0^ and *Q*^1^. The model error at this step is given by

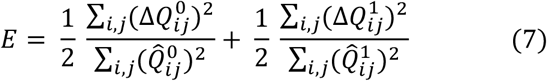

where the difference matrices 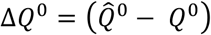 and 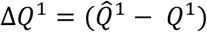 are normalized by empirical covariances. The desired Jacobian update is the matrix:

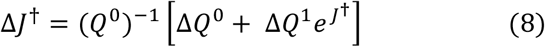

Which decreases the model error *E* at each optimization step as in a gradient descent. The connectivity update is

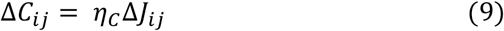

for existing links only; other weights are forced at 0. Here, *η*_*C*_ refers to rate of change of connectivity which was set to 1E-4. We also impose non-negativity for the EC values during the optimization. The best fit and hence the estimated EC corresponds to the minimum of *E*.

### Classification

We used values of EC and FC as measures to classify the cohort in two schemes: (1) Healthy controls vs. patients (2) Patients without a deficit in any of the seven behavioral domains, those with deficit in a single domain, and those with deficits in multiple domains. Classification was done for all participants who underwent resting state scans at each of the three timepoints. For the first classification, we pooled data from healthy controls at two time points separated by three months to improve the balance between class frequencies.

From the EC and FC matrices for each partipant we selected values for existing links in the HCP-SCT (N=4218), and rearranged them in a vector form. Since both the HCP-SCT and FC were symmetric, we used only half of all existing links (N=2109) for classifications involving FC. We then z-scored the values of EC and FC for each participant using the mean and standard deviation of EC and FC across all links within that participant. These z-scored values were the inputs to the classifier which means that the classification relies on the ranking of them rather than the absolute values themselves. We used multinomial logistic regression (MLR), which reduces to logistic regression in the case of binary classification, as a classifier. For each classification scheme, we divided the participants in a random split of training (80%) and test (20%) sets stratified over the classes (i.e. preserving balanced ratios of elements from each class in the training and the test set). MLR classifier using measure values for all links was trained using the training set and its accuracy was evaluated using the test set, thereby assessing the classifier’s ability to generalize to new samples. The procedure was repeated for 100 splits and a mean accuracy across splits was calculated. The significance of this value, at 0.05 level, was tested against the null hypothesis of it arising by chance by comparing it with a distribution of chance-level accuracy values. Here we randomly permuted class labels across individuals and split the data 20 times into train and test splits and obtained the test-set accuracy in each case. This was repeated for 100 random permutations of class labels and the accuracy values were pooled together to construct the distribution of chance-level accuracy values. Each permution preserved the relative proportion of classes. This allowed us to identify the measures (either EC or FC) which showed significantly higher mean accuracy than the chance level for each classification. We also calculated the confusion matrices for each classification that compares the true class labels with those predicted by the classifier (Gilson *et al*., 2019).

### Identification of important links for classification

Here we sought, for each measure (EC or FC) that performed significantly better than the chance-level, the most important links for that classification using recursive feature elimination (RFE). Here, we used data from 90% of participants in each class as the training set and the remaining 10% as the test set. We ranked the links via the RFE using the training set, trained the MLR classifier on it using only the highest ranked link at first, and calculated its accuracy on the test set. We progressively added one link at a time, in the order of the ranking, and calculated the accuracy curve as a function of all links. We then identified, as the predictive links for that train-test split, a minimum number of highest ranked links for which the slope of the smoothed accuracy curve was less than 1E-6 (to locate saturation) and the accuracy was at least 90% of the mean accuracy obtained with all links considered. We repeated this procedure for 90 train-test splits and links that were part of the predictive set in at least 20% of all splits constituted the most-predictive set. This percentage was determined by recalculating the classification accuracy using the most-predictive set of links at different percentage thresholds. As Figures S2 and S3 show, the accuracy begins to drop below 100% at a threshold of 20% and continues to drop for higher thresholds. As the threshold increases beyond 20%, the most predictive sets contain less links that do not sufficiently represent different train-test splits and hence the accuracy drops. We also pooled the predictive links from all splits and compared the size of the pooled set with the total number of links to test the stability of the ranking.

The rank of links in each training set was linearly transformed to a normalized rank so that the highest ranked link was rank-normalized to 1 while the lowest one was rank-normalized to zero. Subsequently, the normalized ranking for each link was averaged across all train-test splits.

## RESULTS

### Effective connectivity estimation for individual participants

We first obtained functional connectivity matrices for each participant using zero-lagged Pearson’s correlation between BOLD signals of all 324 ROIs. Then we fit the model to reproduce the zero-lagged and lagged empirical BOLD covariance matrices by estimating EC for a mask of plausible neuro-anatomical links. As mentioned in the methods section, this mask, termed EC_mask_Ctl (Figure 1D), was identical for each healthy participant. It included all links with value stronger than 0.5% of the strongest SC value in SC-Ctl (Figure 1A), which was a SC averaged across all healthy individuals from this study, and all links in the HCP-SCT (Figure 1B) which was a template created from a publicly available tractography atlas (Yeh *et al*., 2018) constructed using DWI data from 842 HCP participants. For each patient, we built an individual mask, EC_mask_Pat_i_ (Figure 1E), by excluding from EC_mask_Ctl, all completely damaged links (Figure 1C).

**Figure 1.**
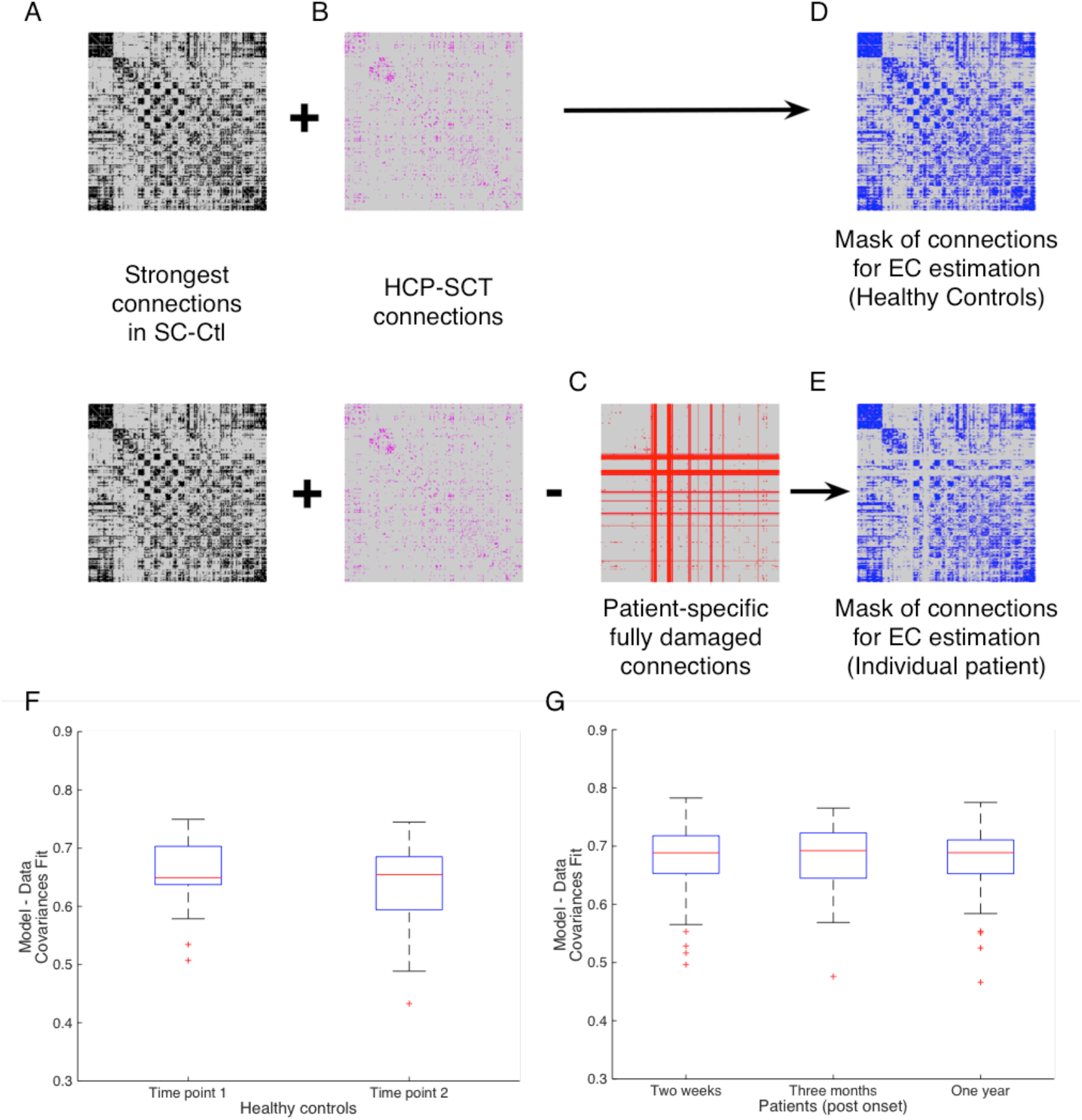
Masks of structural links used in estimating the effective connectivity for healthy control (D) and each individual patient (E). For healthy controls, the mask is the union of (A) links stronger than 0.5% of the strongest value in the SC_Ctl, matrix averaged across healthy control participants (SC_Ctl, see methods), and (B) existing links in the HCP-SCT. From this mask we remove all links that are completely damaged by the lesion in an individual patient (C), to find an individualized mask for the patient. F, G: Box plots of Pearson correlation values between the empirical and model lagged and non-lagged co-variances for (F) age and education matched healthy controls at two time points, three months apart and (G) stroke patients at three time points: 2 weeks, 3 months and 1 year post onset of stroke.

We included both the strongest links in SC-Ctl as well as those present in HCP-SCT because while the links in the HCP-SCT were very reliable they were too few (N=4218) to obtain a robust estimate of EC for reproducing empirical covariances. Hence it was necessary to include the strongest links in the SC-Ctl (N=28256) in the estimation of EC. However, sparsity and reliability of links in the HCP-SCT allowed us to use only them for classification and biomarker identification, thereby keeping the computational cost low without losing the prediction accurary (Figure S1). More importantly, we incorporated the impact of lesions on white matter pathways in each patient using the corresponding structural disconnectome obtained by identifying streamlines in the HCP-SCT that passed through the lesion.

Using the structural connections mask for each participant as a constraint on the model (akin to a topological prior), we inferred individual EC matrix that maximized the Pearson’s correlation between model and empirical lagged and non-lagged covariances. Average fit across healthy controls and patients was found to be 0.65 and 0.7 respectively which corresponds to a statistical *R*^*2*^ of 0.4-0.5 (Figure 1F-G).

### Distinction of patients from healthy controls using EC and FC

Next we turned our attention to assess the predictive power of EC and FC for different classifications of this cohort. As mentioned earlier, values of EC and FC for only the links present in the HCP-SCT were used to classify. We pooled together the measures from two time points for healthy controls together, thus we had EC and FC values for 45 healthy control scans, and, 95, 73, and 53 patients at the two-weeks, three-month, and one-year time points respectively. Top panels of Figure 2 (A-C) show the distributions of test-set accuracy of these measures in classifying healthy controls versus patients, at the three time points respectively, in comparison with the corresponding chance-level accuracy distributions. Here, we pooled the EC and FC values from the scans of healthy controls at two time points together to improve the balance of number of participants in each class. Mean classification accuracy for both EC and FC was higher than the chance level accuracy values at the significance level of 0.05. However, EC performed significantly better than FC (p < 0.001; unpaired t-test, Bonferroni corrected for 3 comparisons) for classification at all three time points. We found the difference in the mean accuracy values had a medium to large effect size (Cohen’s d = 0.46, 0.98 and 0.71 at the three time points respectively). The accuracy at the one-year time point was comparatively lower than the first two time points reflecting the fact that some patients recovered from stroke and their connectivity became again similar to those of control participants. Bottom panels of Figure 2 display the confusion matrices for the three time points using EC (D-F) and FC (G-I) values. The asymmetry of confusion matrices at the two-weeks and three months time points shows that classification error is mostly due to some patients being classified as controls. Weighted precision scores, averaged across train-test splits, with EC (0.86, 0.85, 0.81 for the three time points respectively) were significantly higher (p < 0.05; unpaired t-test, Bonferroni corrected for 3 comparisons) than the corresponding mean scores (0.83, 0.78 and 0.75) with FC.

**Figure 2.**
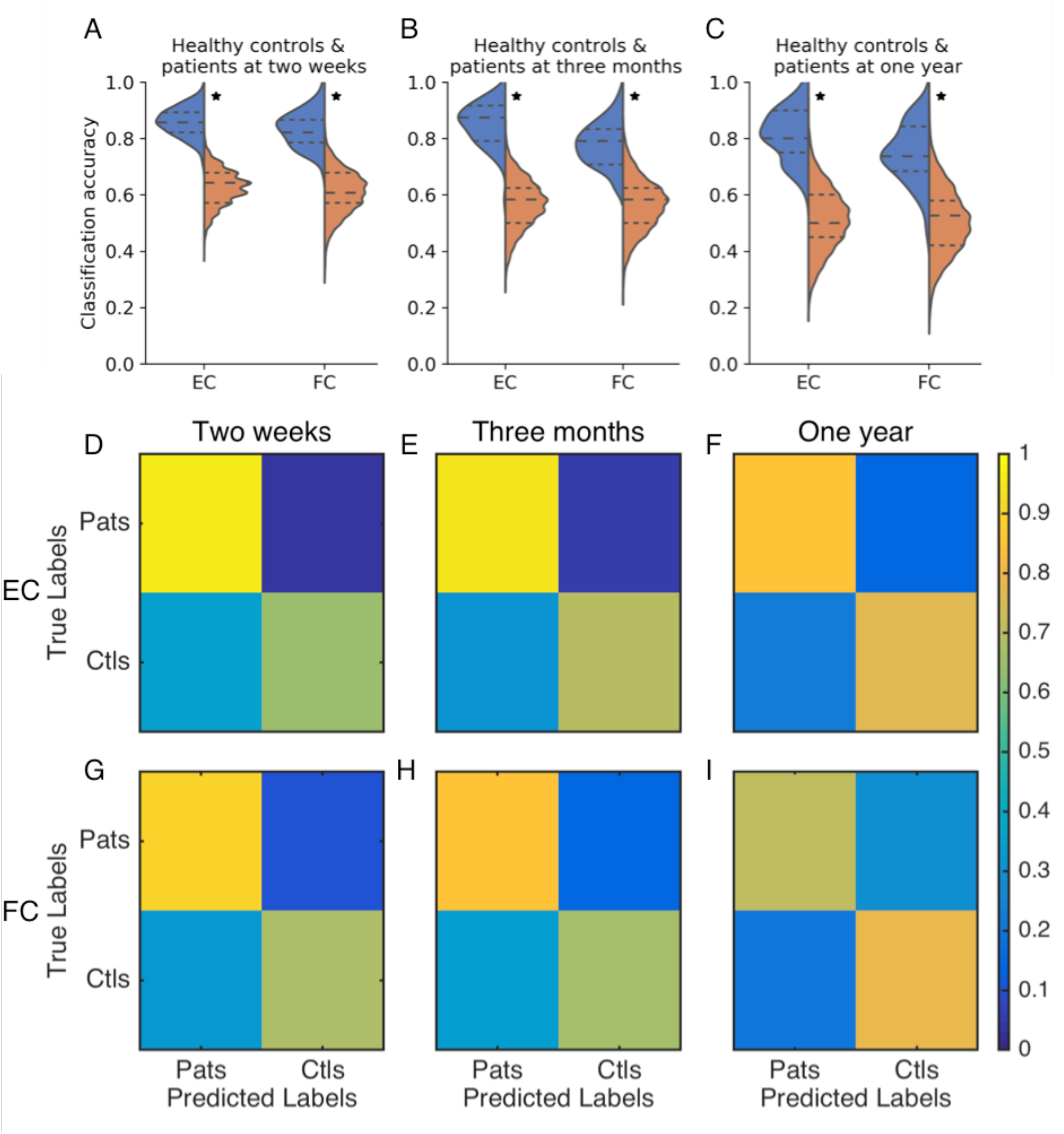
Distributions of accuracy in distinguishing healthy controls and patients using EC and FC values for all links in the HCP-SCT at two weeks (A), three months (B) and one year (C) post onset of stroke. Orange violin displays chance-level accuracy distribution and asterix denotes significantly higher mean accuracy than chance level (p < 0.05). Mean EC accuracy was significantly greater than mean FC accuracy at all three time points (p < 0.001, unpaired t-test, Bonferroni corrected for 3 comparisons). D-I: confusion matrices using EC (D-F) and FC (G-I) values for classifying healthy controls and patients at the two weeks (D, G), three months (E, H) and one year time point (F, I).

Mean accuracy obtained using FC values for all links between 324 ROIs was not significantly different from that obtained using FC values for only the links in the HCP-SCT (Figure S1), confirming that HCP-SCT links were sufficiently informative for classification.

### Identification of EC and FC biomarkers for distinguishing stroke patients from healthy controls

Since mean classification accuracy using EC and FC values for all links in the HCP-SCT was found to be significantly higher than the chance level accuracy values, we sought to extract EC and FC based biomarkers for this classification. A biomarker in this context is a subset of all existing links in the HCP-SCT that is sufficient for an accurate classification. Here, we used recursive feature elimination (RFE) to first rank the links using a training set (90% participants) and then identified a minimum number of most predictive links with which the classification accuracy saturated at a sufficient level (see methods section for details). This was repeated for 90 train-test splits. We tested whether this ranking of links was robust across train-test splits by pooling together the predictive links that featured in at least 2 splits. The number of links in this union was found to be only a small fraction of all links - 9.6%, 12.7% and 7.2%-, in case of EC, and a similarly small fraction - 18.2%, 4.6% and 10.3% -, in case of FC at the three time points respectively, implying that the ranking of links was not specific to the part of the dataset used to train the classifier & was indeed consistent across all train-test splits. Finally, we chose the links that were found to be predictive in at least 20% of all splits as the biomarkers. As described in the methods, this threshold was chosen because the classifier accuracy with only the most predictive links started dropping below 1 beyond this percentage threshold (Figure S2, S3) in most cases.

Figure 3A-C display the biomarker EC links for the three time points respectively while Figure 3G-I display the corresponding, most predictive FC links. Figure 4 demonstrates average ranking of all predictive links, with EC (Figure 4A-C) and FC (Figure 4D-F) as measures, belonging to each within- and inter-RSN (within & inter-hemispheric) block to the classification at the three time points respectively. The asymmetry in the average ranking of predictive RSN blocks highlights the asymmetry and directionality in the EC estimation. Therefore, we compared, for each RSN, the average ranking weighted by the fraction of links in each of the four sub-groups, namely, (a) efferent, inter-hemispheric (IH), (b) afferent, IH, (c) efferent, intra- or within-hemispheric (WH) and (d) afferent, WH, of predictive EC links. We also found average weighed ranking across all WH and IH predictive links for classifications involving FC. Figure S4 shows mean (+/-SEM) normalized rank across splits of these sub-groups of links for healthy controls versus patients classification at each time point. Firstly, the IH links were relatively more important than the WH links for most RSNs for classification at all three time points. At the two-weeks time point, efferent WH links from DAN and VAN, IH links from DMN and SMH and afferent WH links to DMN and IH links to SMH were significantly more important than other groups for EC-based classification while for FC-based classification, WH links in VIS, VAN, DAN and DMN, and IH links in SMH and COP were most important. At the three-month time point, most relevant contributors to EC-based classification were efferent WH links from VAN and IH links from COP, SMH, VAN, DMN and VIS; afferent WH links to SAL, DMN and SMH and IH links to SMH while for FC-based classification, COP and DAN WH links and VIS and COP IH links were the most important. Finally for the one-year EC-based classification, efferent WH links from SMM and VAN and IH links from DMN; afferent WH and IH links to SMH, and for the FC-based classification, WH COP links and IH links in COP, VIS, SMH, AUD and VAN were the most prominent contributors. Here, the mean rankings were compared across groups by performing one-way Kruskalwallis test for each type of links (efferent/afferent, WH/IH) followed by post-hoc tests corrected for multiple comparisons.

**Figure 3.**
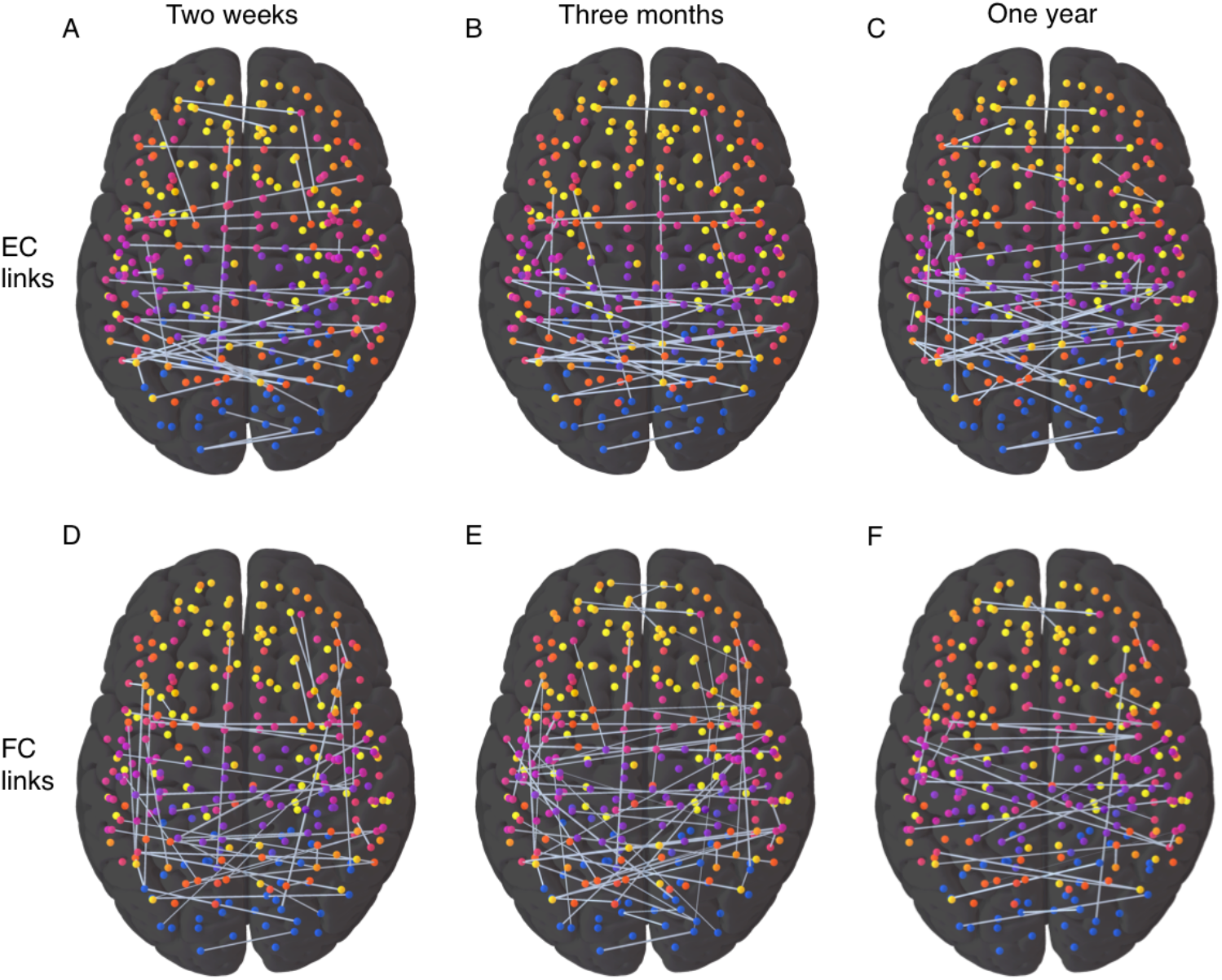
Predictive links using EC values (top panels) and FC values (bottom panels) for classification of healthy controls and patients at the two weeks (A, D), three months (B, E) and one year (C, F) time point respectively. The node colors represent different RSNs.

**Figure 4.**
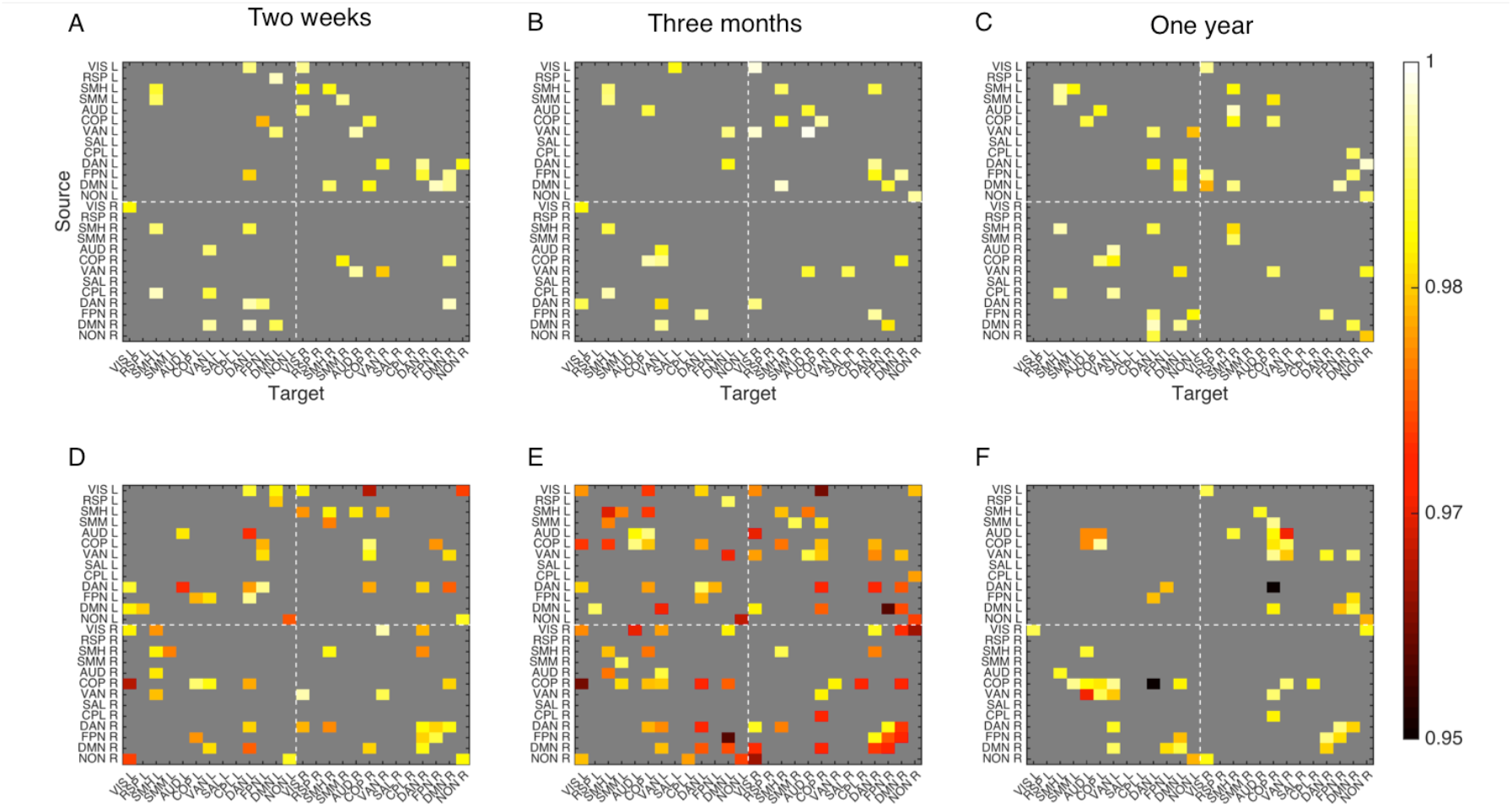
Predictive RSNs for EC (top panels) and FC (bottom panels) for the classification of healthy controls versus stroke patients at each time point (2 weeks, A&D; 3 months B&E; 1 year C&F). White dashed lines divide the RSNs by hemispheres (top left: left hemisphere; bottom right: right hemisphere). As EC is asymmetric, the top matrices also show the between-RSN as well as between hemisphere directionality of predictive links.

We compared the normalized ranking of all links, averaged across train-test splits, at one time point with other time points for both EC and FC as well as between EC and FC at each time point. As Figure S5 shows, we found statistically significant Pearson’s correlation in all cases, however, the sets of predictive links in any two cases had few common members. This points to the fact that physiological changes caused by stroke are not static and while changes at earlier time point could recover at later time points, network re-organizations could happen at later time points.

We also compared the average normalized ranking of all and most predictive EC and FC links, at each time point, with their structural disconnections averaged across patients. Note that while the completely damaged links from the HCP-SCT in each individual patient were not included in the estimation of individual EC, they were included (with their EC value set to zero for the corresponding participant) in classification as well as in the identification of predictive links. As Figure S6 shows, we did not find any significant correlation between these two measures, indicating that strength, across participants, of structural disconnection of a link alone did not explain its importance for the classification at any time point. We also did not find any correlation between the normalized rank and frequency of disconnection of all or the most predictive EC and FC links. Thus information about changes in the resting state of patients carried by predictive links is not trivially explained by structural impact of stroke.

### Predicting number of behavioral deficits & prognosis

Next, we considered, in stroke patients alone, the classification of behavioral deficit severity based on the number of impaired domains. We calculated the z-scores, using the mean and standard deviation of healthy controls, from the seven-factor scores that account for the most behavioral variance across the different neuropsychological test scores in four domains – language, motor, attention, and memory. Z-scores of −2 or less defined a deficit in a domain. We then divided patients, at each time point, into three classes: (a) patients without a deficit across all seven factors, (b) patients with a deficit in one factor, and, (c) patients with deficits in multiple factors. At time point 1, 16 patients showed no deficit, 30 with a single domain deficit and 49 with multi-domain deficit. At time point 2, 23, 22 and 25 patients and at time point 3, 20, 25 and 9 were in these three classes respectively. Then, we used EC and FC links to classify patients in these three classes. In the first analysis, EC and FC links at one time-point classified patients’ number of deficits at the same time-point. In a different and clinically more pertinent analysis from a prognostic point-of-view, we used links at the two weeks time point to predict patients’ number of deficits at later time points.

Figures 5A-C show the accuracy of same time point EC and FC links in classifying patients’ number of deficits at the two weeks, 3-month, and one-year time points. Figures 5D-E show the prognostic accuracy of two-weeks time point EC and FC links in classifying patients at the three months and one-year time points respectively. FC did not perform better than the chance level for any of these cases while EC at two weeks time point was significantly more accurate than the chance level accuracy values in classifying patients at both two weeks as well as one-year time points. Unsurprisingly, the mean accuracy of two weeks EC links in classification of two weeks and one year time points patients was significantly higher than the accuracy of two weeks FC links (p < 0.05, unpaired t-test, Bonferroni corrected). Here, again, we found a medium to large effect size (Cohen’s d = 0.50 and 0.72 at the two time points respectively) difference in the mean accuracy of EC and FC.

**Figure 5.**
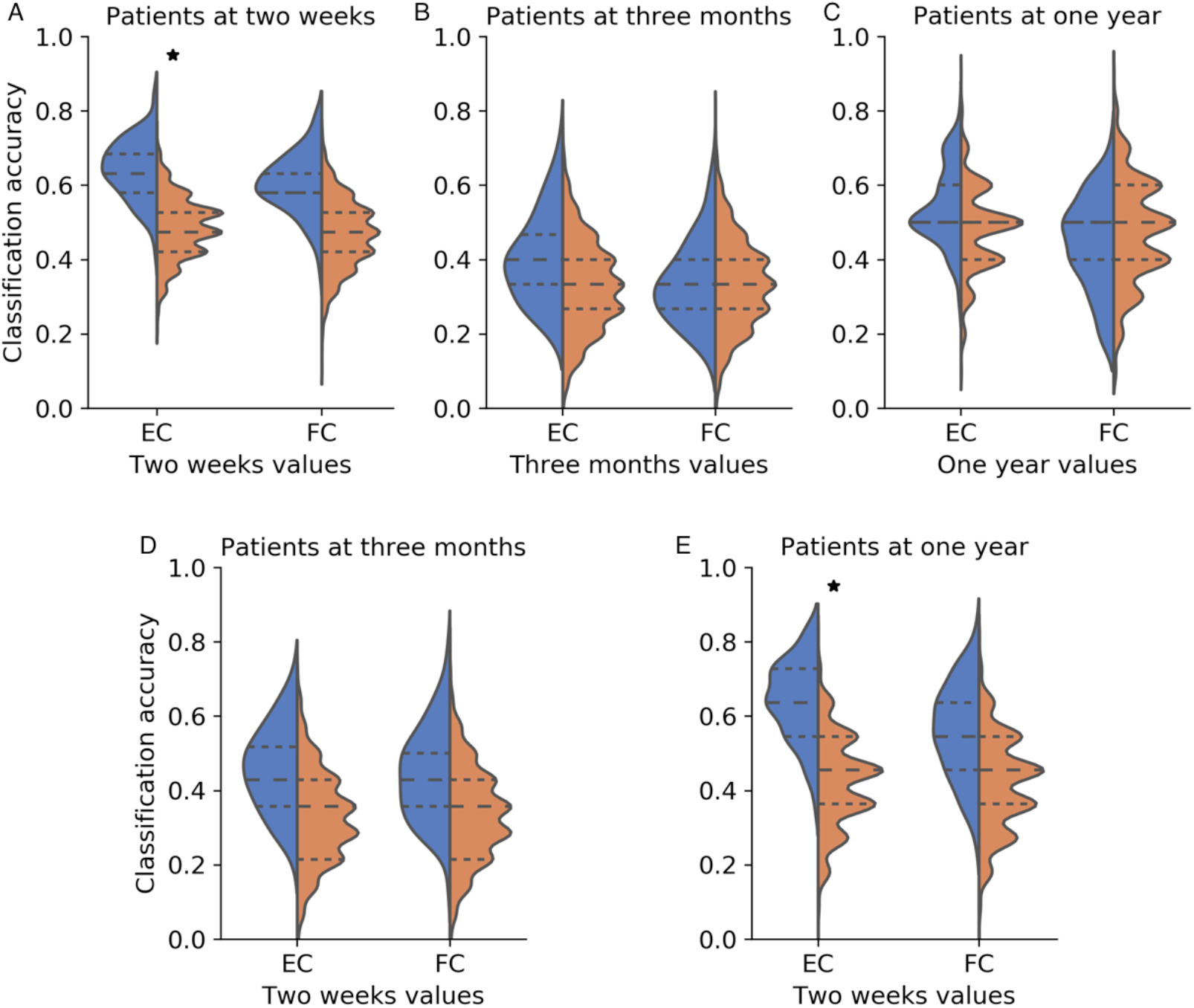
Accuracy of classification of stroke patients according to the number of deficits using EC and FC values of all links in the HCP-SCT. Top panels: classification of patients at the two weeks (A), three months (B) and one year (C) time points using EC and FC values at the same respective time points; bottom panels: prognostic accuracy of EC & FC -- classification of patients at the three months (D) and one year (E) time points using EC and FC values at the two weeks time point. Orange violin displays chance-level accuracy distribution and asterix denotes significantly higher mean accuracy than chance level (p < 0.05). Mean classification accuracy using two weeks EC links for classifying patients at two weeks and one year time points was significantly higher than that using corresponding FC values (p < 0.01, unpaired t-test, Bonferroni corrected for five comparisons)

Figure 6 displays the confusion matrices for classification of patients at all three time points using two weeks EC (A-C) and FC (D-F) links. Weighted precision scores, averaged across train-test splits, for classification with EC links were 0.64, 0.46 and 0.64 and with FC links were 0.60, 0.44 and 0.56 for the two weeks, three months and one-year time points respectively with the difference being statistically significant for the two weeks and one year time point (p< 0.05). While both EC and FC predict the largest class of patients with deficits in multiple domains at the two weeks time point (Fig 6 A, D) well, both do not predict participants without any deficit well. At the one-year time point, since several patients with multiple deficits at the two weeks time point have recovered, this is a very small class (9 participants) and both EC and FC fail to predict it well. Additionally, at the one-year time point, we combined both classes of patients with deficits to make the class-sizes more comparable. Both two-week EC and FC performed this binary classification (no deficits versus deficit) at the one-year time point accurately (Figure S7A), however, while FC predicted the no-deficit class better, EC predicted the deficit class more accurately (Figure S7B-C.

**Figure 6.**
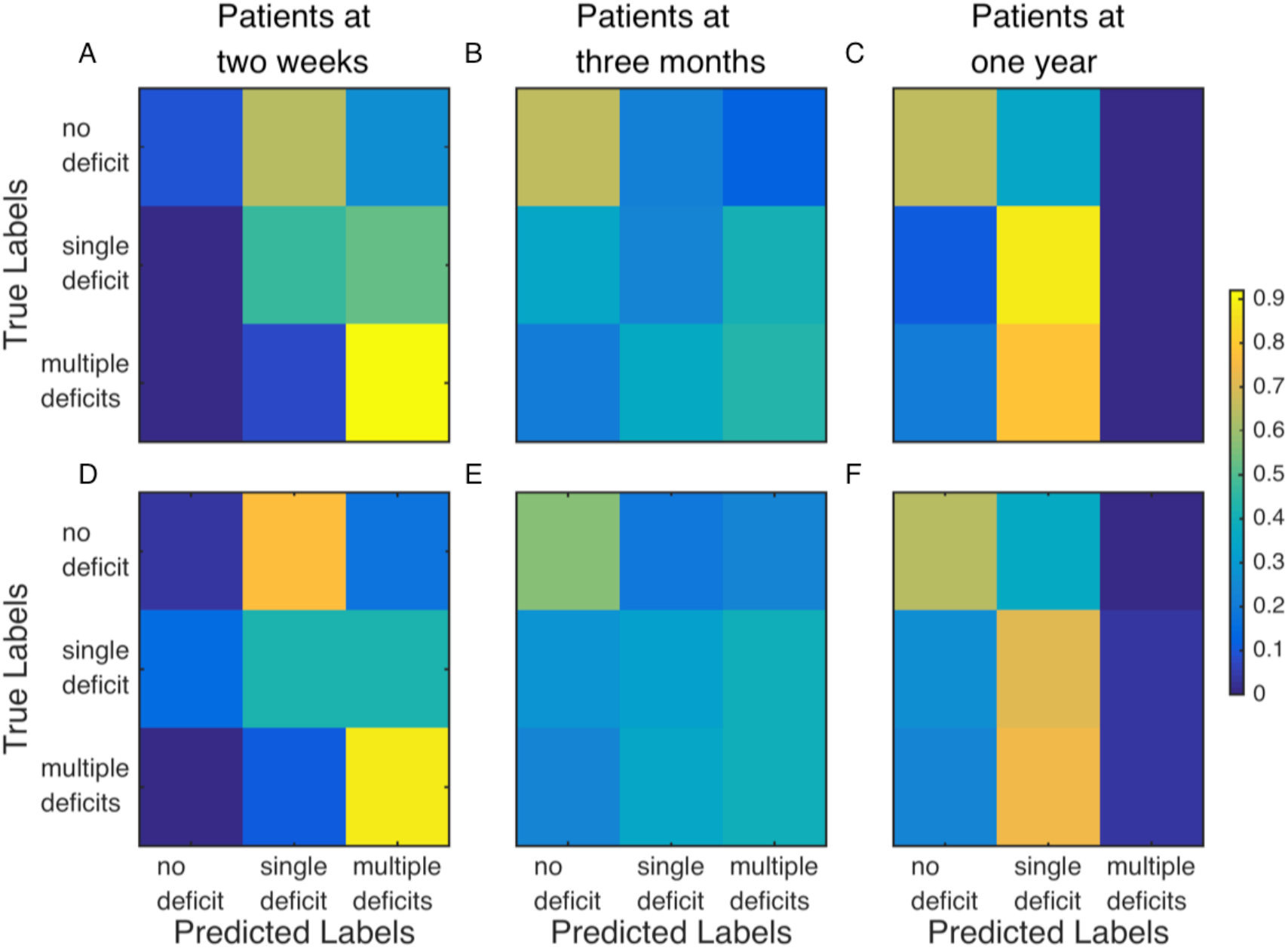
Confusion matrices for the three-way classification of patients by number of behavioral deficits. Only two weeks EC (top row) and FC (bottom row) values for all links in the HCP-SCT are used for classifying patients at the two weeks (A, D), three months (B, E), and one year (C, F) time points.

### Predictive EC links for number of behavioral deficits and prognosis

Next, we looked for the biomarker links whose two weeks EC values are most predictive of patient deficits at two weeks and one-year time points. The ranking of links across different training sets for this classification at both the two weeks and one-year time point was robust as Figures S8A-B show. The fraction of all EC links that were predictive in at least 2 of the 90 realizations for this classification at the two weeks and one-year time point were 11% and 9.7% respectively. The normalized ranking of all links, averaged across all training sets, for classification of participants at the two time points was correlated (r=0.32, Figure S8C), however, the most predictive links in the two cases had few common members. Lastly, the average normalized ranking of all and predictive links did not correlate with the average structural disconnection values for these links (Figures S8D-E) as was also the case with the classification of healthy controls versus patients. The ranking of all and the most predictive links did not correlate with the frequency of structural disconnection for these links either.

Figure 7A-B display these predictive EC links while Figures 7C-D display the average ranking of each within and inter-RSN (within & inter-hemispheric) block to the classification of patients at the two weeks and one-year time points. Overall, the afferent links to RSNs (below the diagonal in matrices in Figures 7C-D) were relatively more important than efferent ones originating from RSNs (above the diagonal in matrices in Figures 7C-D) especially for classifying patients at the two weeks time point. Figure S9 delineates the relative importance of efferent and afferent, IH & WH links in predicting patient’s number of deficits at two weeks and one-year time points. Bidirectional WH links in DAN and IH links from and to visual, SMH along with efferent, IH links from DAN and VAN and afferent, IH links to DMN stood out for the two-weeks time point classification. On the other hand, at the one-year time point, efferent WH links from SMH, FPN and DMN and IH links from SMH and DAN while afferent WH links to AUD, DAN & VAN and IH links to VIS, SMH & DMN found to be the most prominent contributors.

**Figure 7.**
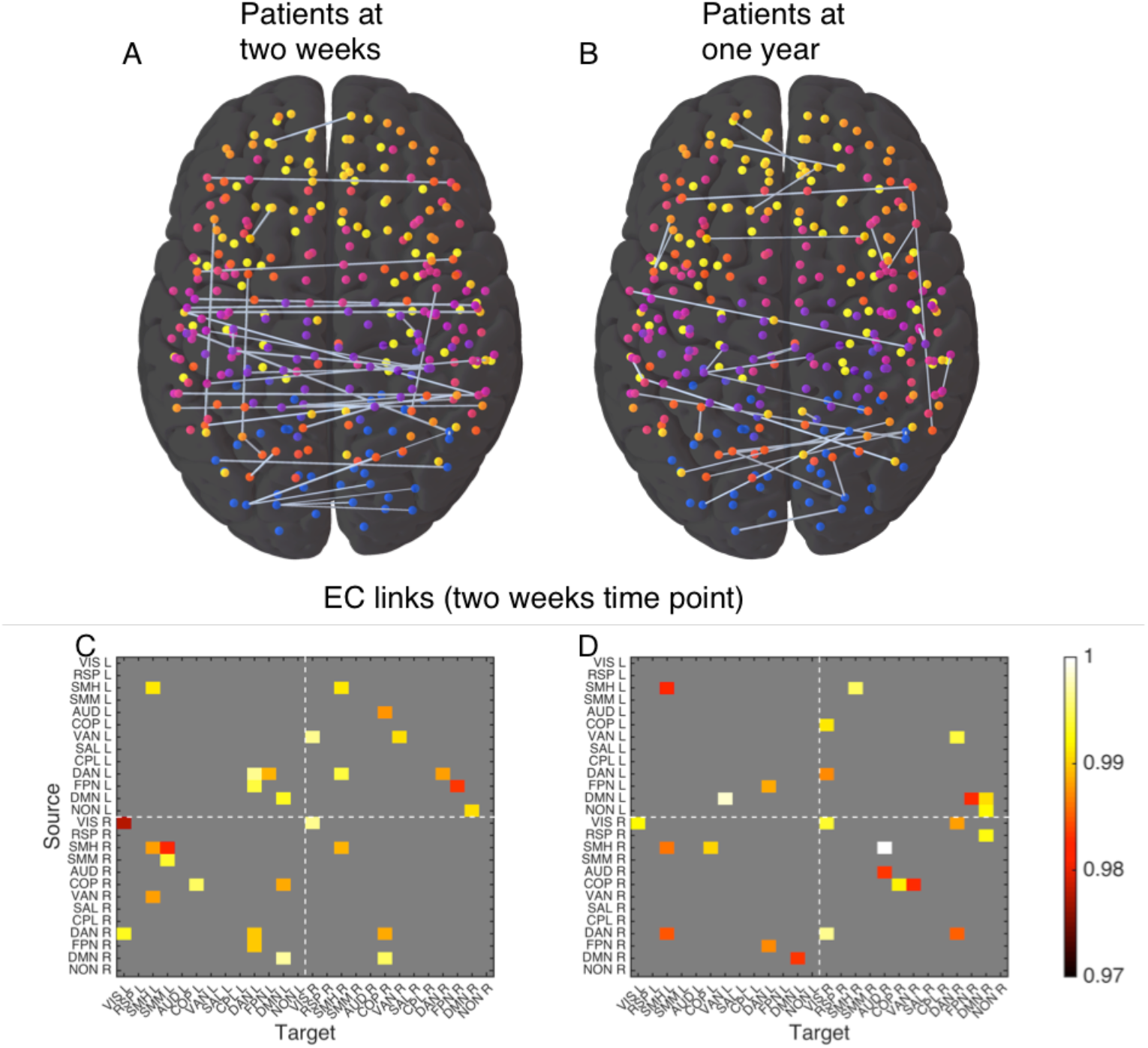
A-B: Predictive links (top panels) and hemispheric RSNs (bottom panels) from EC values at the two weeks time point in classifying patients according to the number of behavioral deficits at the two weeks (A, C) and one year (B, D) time points.

## DISCUSSION

In this paper, we aimed at classifying patients of stroke using resting state functional connectivity and a linear model-based effective connectivity inferred from lagged and non-lagged second order statistics of the resting state fMRI data. At first, we distinguished patients from healthy controls and found that while both EC and FC performed significantly better than the chance level at this task at two weeks, three months and one year post onset of stroke, EC’s mean accuracy was significantly higher than that of FC at each time point. More importantly, we sought to classify patients according to the number of domains in which they showed deficits in comparison with healthy participants at all three time points. Here, mean accuracy of only the EC values at two weeks was significantly more predictive than the chance level of behavioral deficits of patients not only at the two weeks time point but also at one-year time point. Thus, the EC could extract information from resting state neurophysiological signals at an early time point that was relevant, not just for the diagnosis of patients but also their prognosis. We also identified biomarker sub-networks for stroke diagnosis and prognosis.

Estimation of EC in an individual participant is robust when a mask of neuro-anatomically plausible links is used as a prior in the inference(Gilson *et al*., 2019). In this study, we combined two SC masks to, first, define the links for which EC was estimated for all healthy individuals and, second, to define the corresponding links for each individual patient by excluding from the combined mask all completely damaged links in that individual’s brain. The first mask, consisting of strongest links in SC-Ctl, the SC averaged across healthy controls obtained using DWI and probabilistic tractography, was necessary to have a reasonable minimum number of estimated links (27%), so that the fit between empirical and model covariances was sufficiently high (∼0.7 on average). The second mask, consisting of existing links in the HCP-SCT, found using end-to-end tractography with DWI scans in 842 participants of the human connectome project, was too sparse (4% edge density) to obtain a sufficiently high fit between model and empirical covariances. However, it was necessary to estimate EC for these links as they were used for classification; its sparsity ensuring low computation cost of biomarker identification. Secondly, and more importantly, FC, averaged across links existing in the HCT-SCT was found to be significantly stronger than the average across the non-existing links (Griffis *et al*., 2020) in both the healthy and the patients groups. The fact that they were informative for classification was evident from the mean levels of accuracy we found in comparison to the corresponding levels when all links were used (Figure S1).

While both EC and FC were significantly more accurate than the chance level at distinguishing patients from healthy controls at all three time points, EC’s performance was better than FC especially at the two later time points as the confusion matrix in Figure 2 also shows. Recent papers on recovery from stroke (Ramsey *et al*., 2016; Siegel *et al*., 2018) have shown that as patients recover from behavioral deficits, a corresponding recovery of FC alterations observed at the sub-acute stage (1-2 weeks post onset) takes place. Therefore, it is reasonable to expect that as patients recover, the magnitude of FC differences between healthy controls and patients at later time points is reduced in comparison with the early time point. Hence, making the distinction between patients and healthy individuals using FC, that only measures spatial correlation, would be less accurate at later time points than at the earlier one. In contrast, the EC, that is inferred using both spatial and temporal correlations, continues to predict the two classes well at later time points.

Predictive power of EC and FC faced a significantly tougher scrutiny in the case of a complex task of predicting classes with varying degree of severity in terms of the number of behavioral deficits. Firstly, the choice of class definitions in this case was driven by the need to balance predictability with interpretability. Sufficient number of participants in each class and, as far as possible, balanced class frequencies were required to ensure predictability. Interpretability was based on clinical considerations. Indeed the notion of behavioral deficit was based on a clinical definition of a score two standard deviations below the average control score. We did consider other class definitions – one, in particular, to assess recovery distinguished patients according to the change in either the within-domain deficit or in the number of deficits across domains. However, here, we did not have sufficient samples in each such class to ensure predictability. Also, some patients recovered from deficit in one domain while worsened in another. So labeling such cases appropriately was a challenge from the point of view of interpretation.

In this classification, FC did not perform better than the chance level. The result did not change whether we considered only the links present in the HCP-SCT or all possible ones. EC values of all HCP-SCT links at two weeks time point, on the other hand, not only predicted patient classes at the same time point with significantly higher accuracy than the chance level, it did so at the one year time point as well. Moreover, as the confusion matrices in Figure 6 demonstrate, the two classes at the one-year time point that were predicted better included patients without any deficit and those with deficit in a single domain. Both these classes include patients that have recovered from their initial deficits. Therefore, the temporal correlation structure found in an early stage resting state fMRI that is captured by the EC is an important contributor to predicting recovery and prognosis of patients.

The objective of this study, apart from comparing classification accuracies of two neurophysiological data driven measures, was also to identify specific links as biomarkers. Most important FC-based signature of patients at all three time points included inter-hemispheric links within several RSNs – namely visual, somato-motor, cingulo opercular, dorsal attention and default mode - and intra-hemispheric links between the dorsal attention, frontoparietal and default mode networks. These observations are in line with the previous research of markers of stroke at the acute/sub-acute stage (Baldassarre *et al*., 2014; Siegel *et al*., 2016). EC biomarkers for the classification at two weeks time point included within-RSN inter-hemispheric links similar to the FC but links to a RSN were not necessarily equally important as links from that RSN. This distinction can be assessed in the case of EC due to its asymmetry. Thus, at two weeks time point, while links in both directions were important for inter-hemispheric links from/to somato-motor, efferent links from DMN were more relevant than afferent ones. Afferent links from somato-motor network were more important at later time points along with efferent links from cingulo-opercular networks (Figure S4).

Since they predicted the number of behavioral deficits in patients at two weeks and one-year time points accurately, we extracted biomarker networks for this classification using only the two-weeks time point EC links. Inter-hemispheric links from and to visual, somato-motor, efferent links from DAN and VAN, and afferent links to DMN were found to be most important for predicting patients’ deficit severity at two weeks post onset of stroke. At one year post onset, when a significant proportion of patients with initial deficit had recovered thus making the no-deficit class the largest, inter-hemispheric links from and to somato-motor along with afferent links to visual network and efferent links from the DAN were most predictive (Figure S9). In case of within-hemispheric links, bidirectional links in DAN were most predictive of patients’ deficit severity at two weeks time point while at the one-year time point, afferent links to DAN, VAN and AUD while efferent links from somato-motor, FPN, and DMN were the two most predictive sets.

In conclusion, our results show that whole-brain model based effective connectivity inferred from second-order statistics of the resting state fMRI data in patients of stroke offers a powerful tool to extract information relevant for clinical prognosis from early stages of the neurological condition. Additionally, longitudinal evaluation of topological signatures that are predictive of patient’s deficit severity allows us to understand changes in the connectivity patterns associated with processes of recovery. Finally, due to the inherent asymmetry embedded in the estimation of EC, these predictive signatures inform on the longitudinal changes in the directionality of interaction between brain networks.

## Data Availability

The full set of neuroimaging and behavioral data are available at http://cnda.wustl.edu/app/template/Login.vm. Specific data and analysis scripts are available on request to the authors.

---

**Figure S1.**
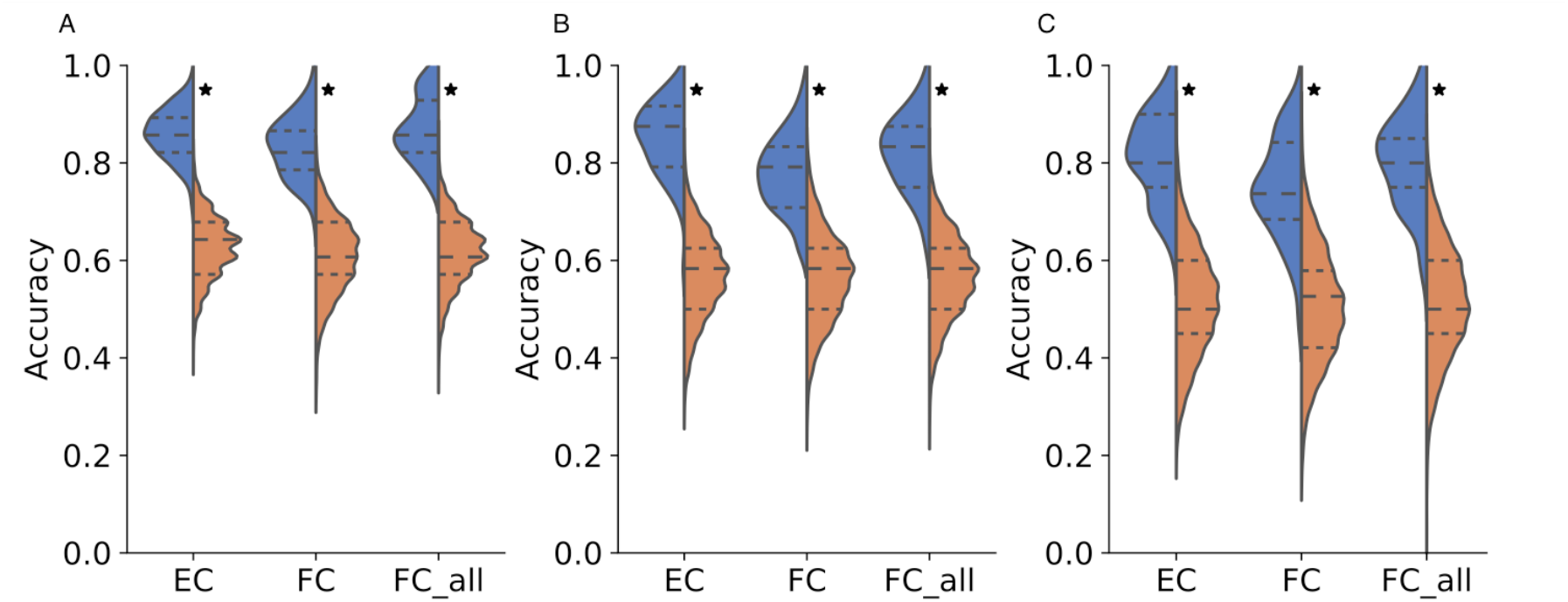
Distributions of test set accuracy values, using EC and FC values for only the links (4218 and 2109 for EC and FC respectively) present in the HCP-SCT (EC and FC) and FC values for all links in the Gordon parcellation (52326 links, FC_all) for distinguishing healthy controls from patients at the two weeks (A), three months (B) and one year (C) time points. Orange violin displays chance-level accuracy distribution and asterix denotes significantly higher mean accuracy than chance level (p < 0.05). Mean accuracy of classification using all FC values is not significantly different than the corresponding accuracy when only a subset, present in the HCP-SCT, are used.

**Figure S2.**
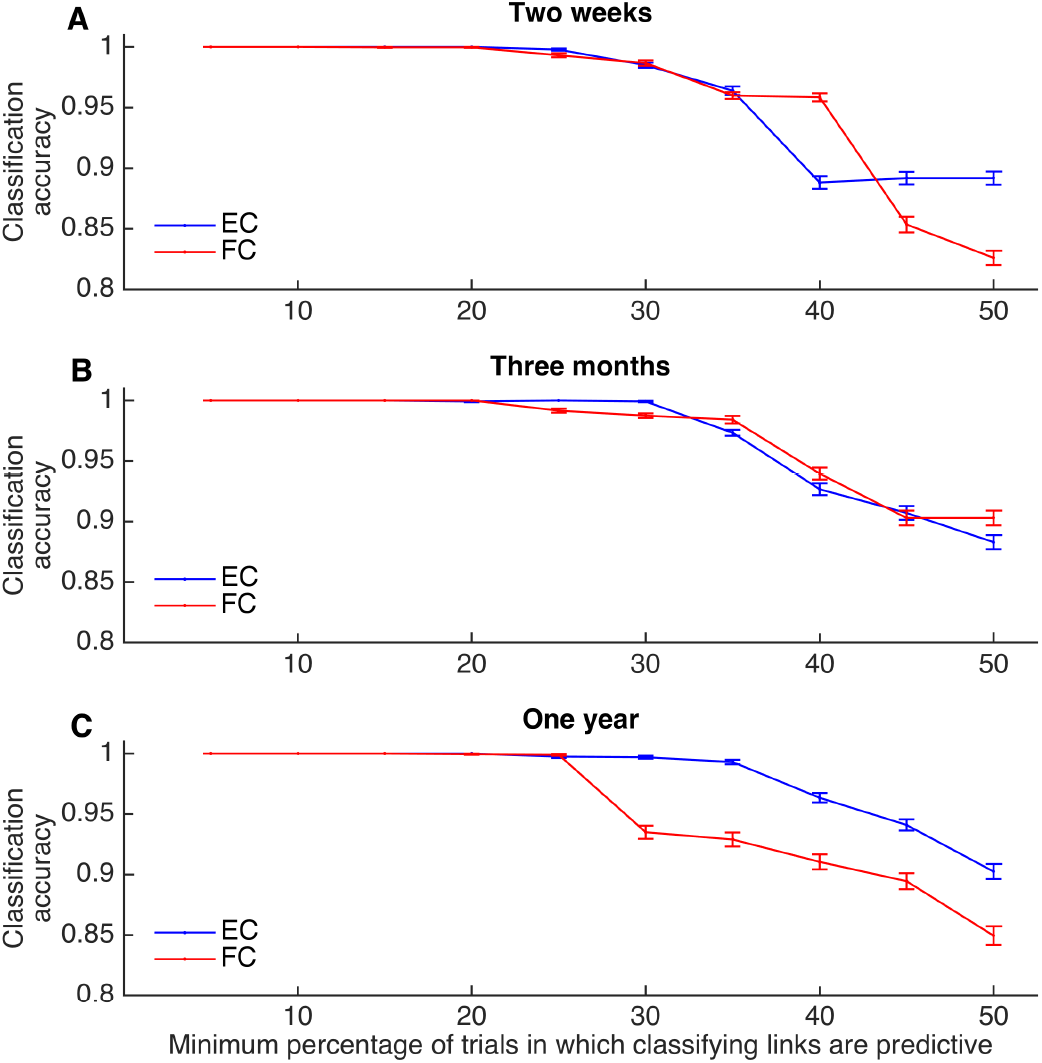
Mean classification accuracy using EC and FC values of only the most predictive links, that are identified as predictive in a range of minimum percentages of train-test splits, for distinguishing healthy controls from patients at the two weeks (A), three months (B) and one year (C) time points. In all cases, the accuracy drops for both measures beyond a threshold of 20%.

**Figure S3.**
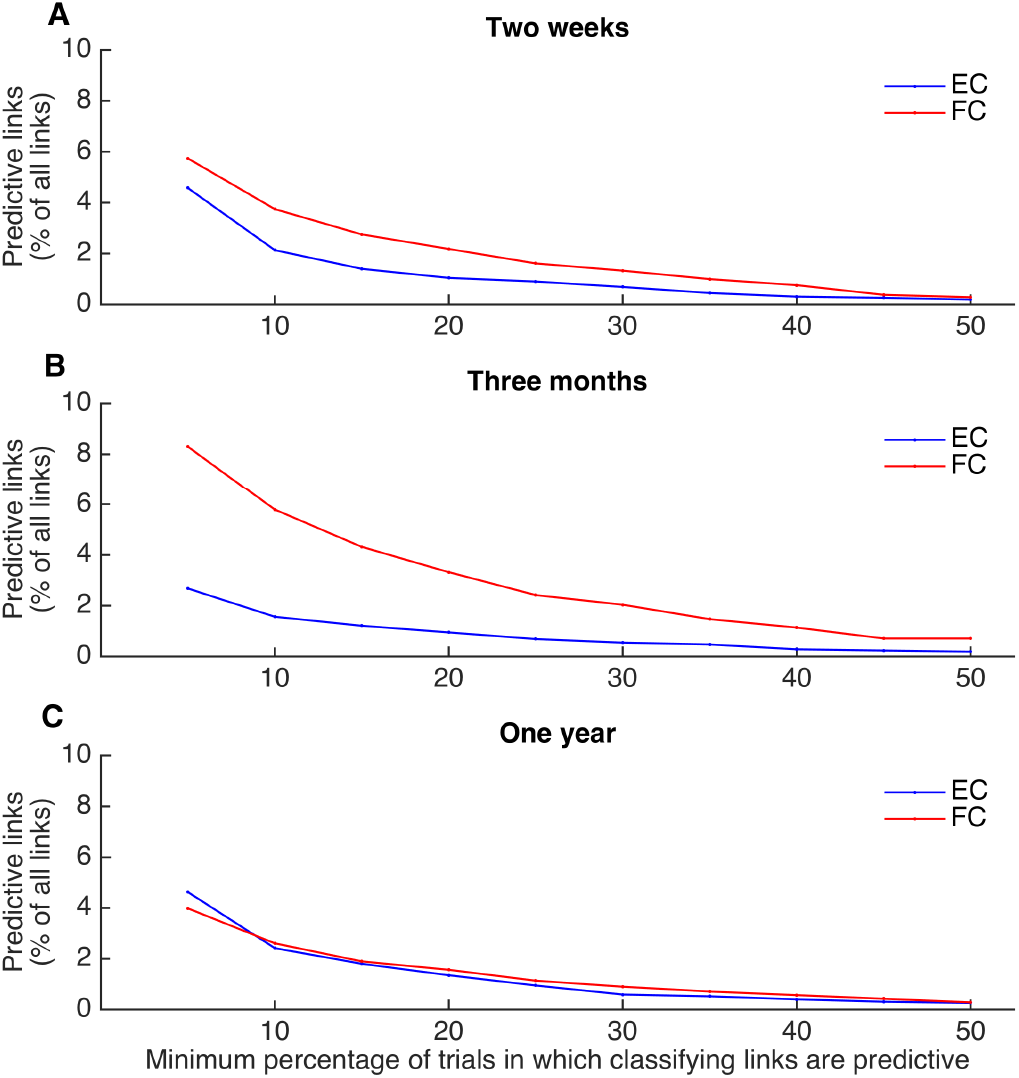
Number of most predictive links as a percentage of all links (N = 4218 for EC and 2109 for FC), that are identified as predictive in a range of minimum percentages of train-test splits, for distinguishing healthy controls from patients at the two weeks (A), three months (B) and one year (C) time points.

**Figure S4.**
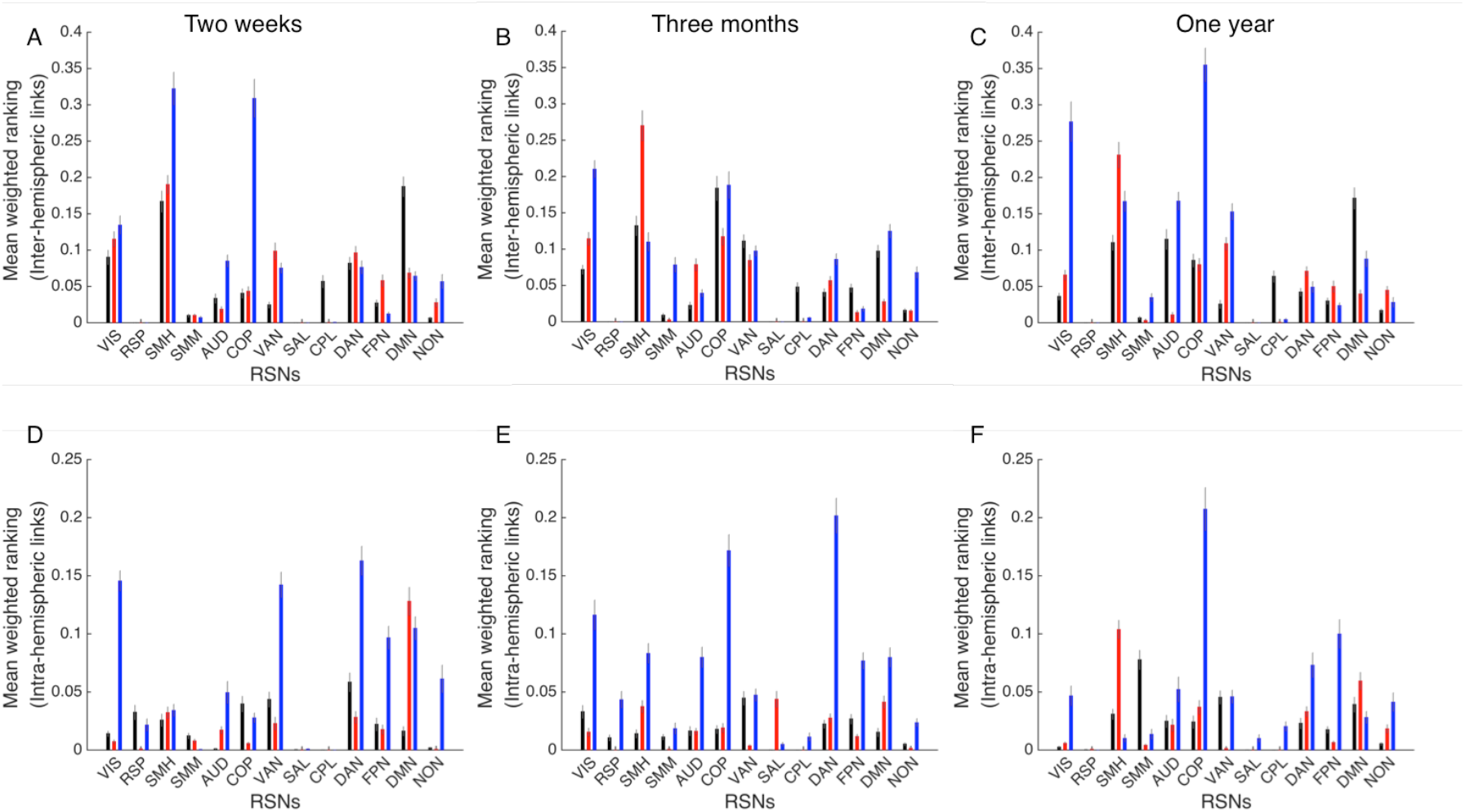
Mean (+/- SEM) weighted ranking of efferent (black) and afferent (red) EC as well as FC (blue) predictive links per RSN for classification of healthy controls and patients at the two weeks (A, D), three months (B, E) and one year (C, F) time points. Top panels consider only the inter-hemispheric links while the bottom panels consider only intra-hemispherical links.

**Figure S5:**
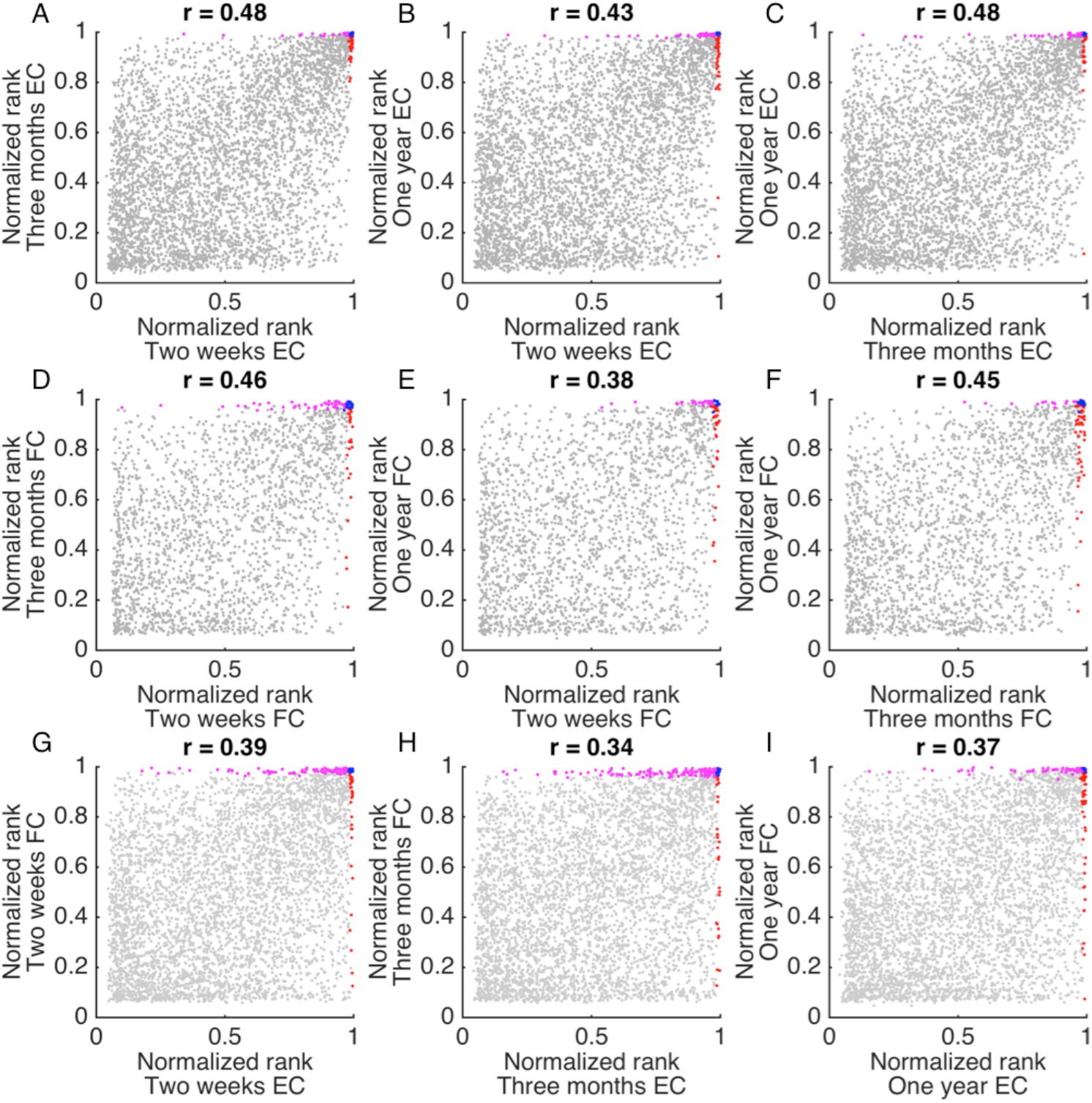
Comparison of normalized ranking, averaged across all train-test splits, of all links present in the HCP-SCT in the classification of healthy controls and patients using their EC (A-C) and FC (D-E) values between different time points and between EC and FC as measures for classification at the same time points (G-I). Pearson’s correlation values (shown in the title of each panel) between rankings of all links are strongly statistically significant (p < 0.001). In each plot, predictive links for the individual measure/time point combination mentioned along x and y axes are displayed in red and magenta respectively while the common ones are shown in blue.

**Figure S6:**
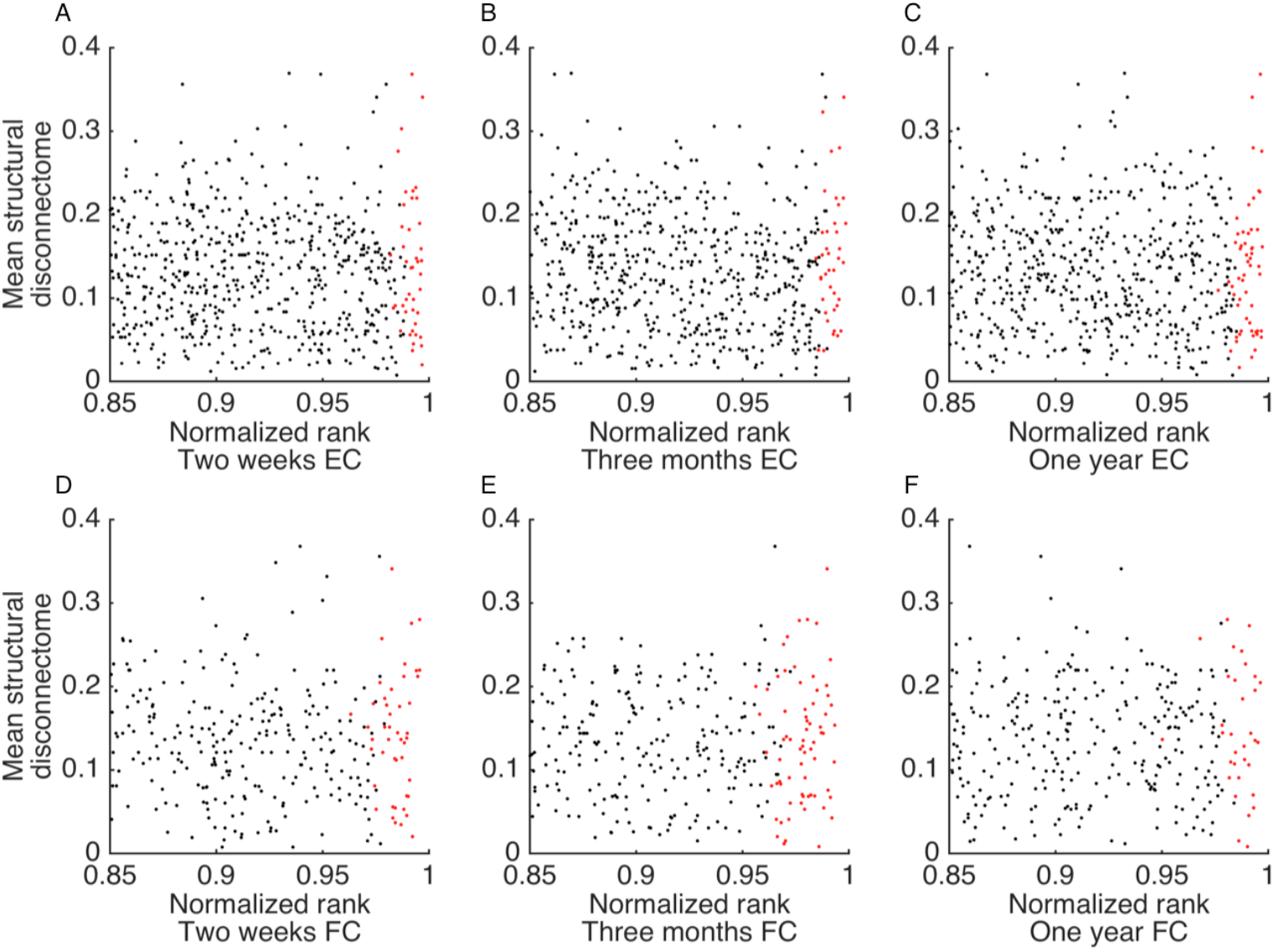
Comparison of normalized ranking, averaged across all train-test splits, for highest ranked links with EC (A-C) and FC (B-E) values used for the classification of healthy controls and patients at the two weeks (A, D), three months (B, E) and one-year (C, F) time points with structural disconnection for each link, averaged across patients. Predictive links in each case are shown in red. In none of the cases, a significant correlation between the ranking and average structural disconnection is found, whether all links in the HCP-SCT or only the predictive links are considered.

**Figure S7.**
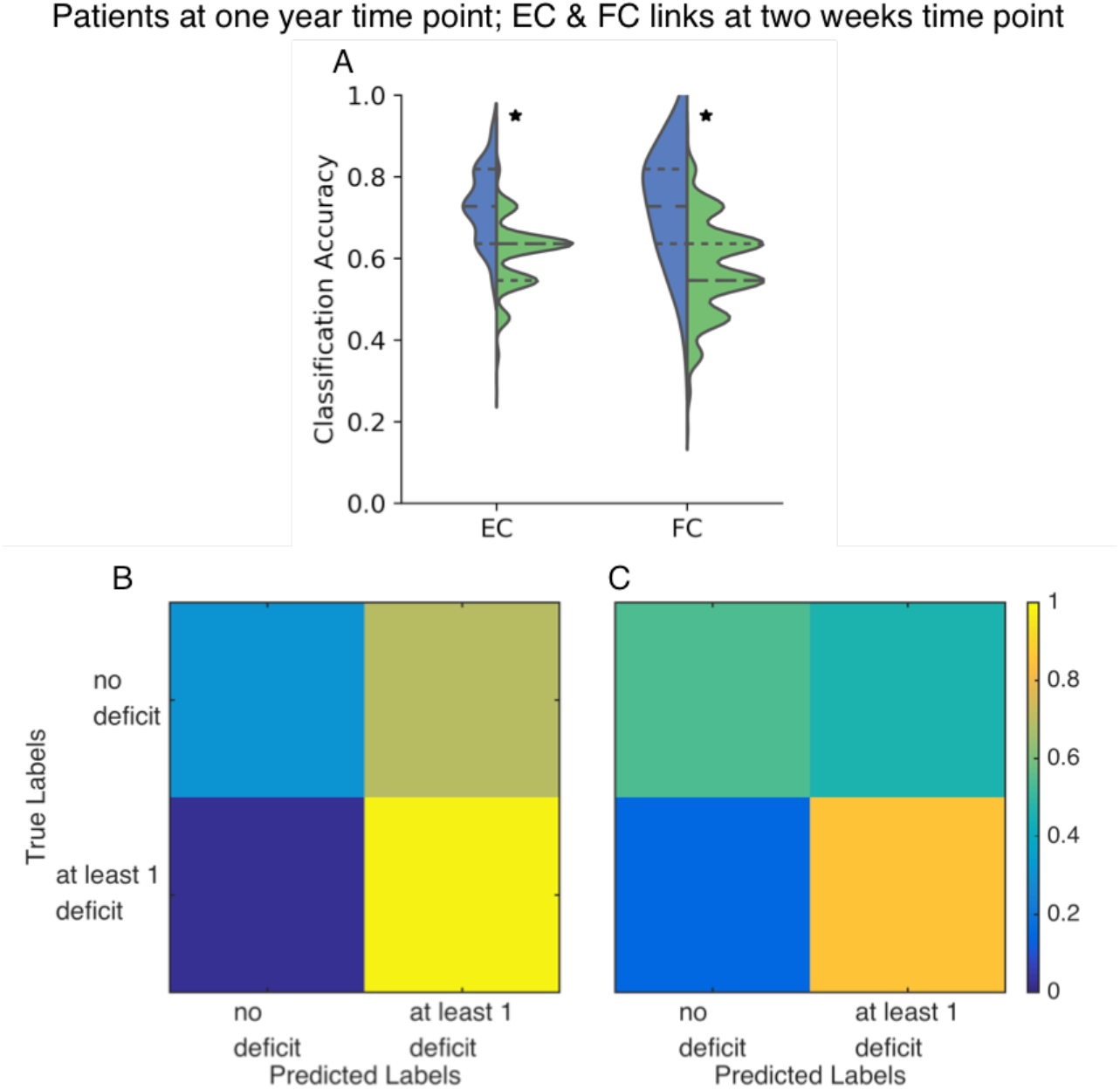
**A:** Distributions of accuracy in distinguishing patients with and without deficit at the one-year time point using EC and FC values for all links in the HCP-SCT at the two-week time point. Asterix denotes significantly higher (p < 0.05) mean accuracy when compared to the chance-level accuracy distribution (green). No significant difference in the mean EC and FC accuracy values was found. **B, C:** confusion matrices using two-week EC (B) and FC (C) values for classifying patients with and without deficit at the one year time point.

**Figure S8:**
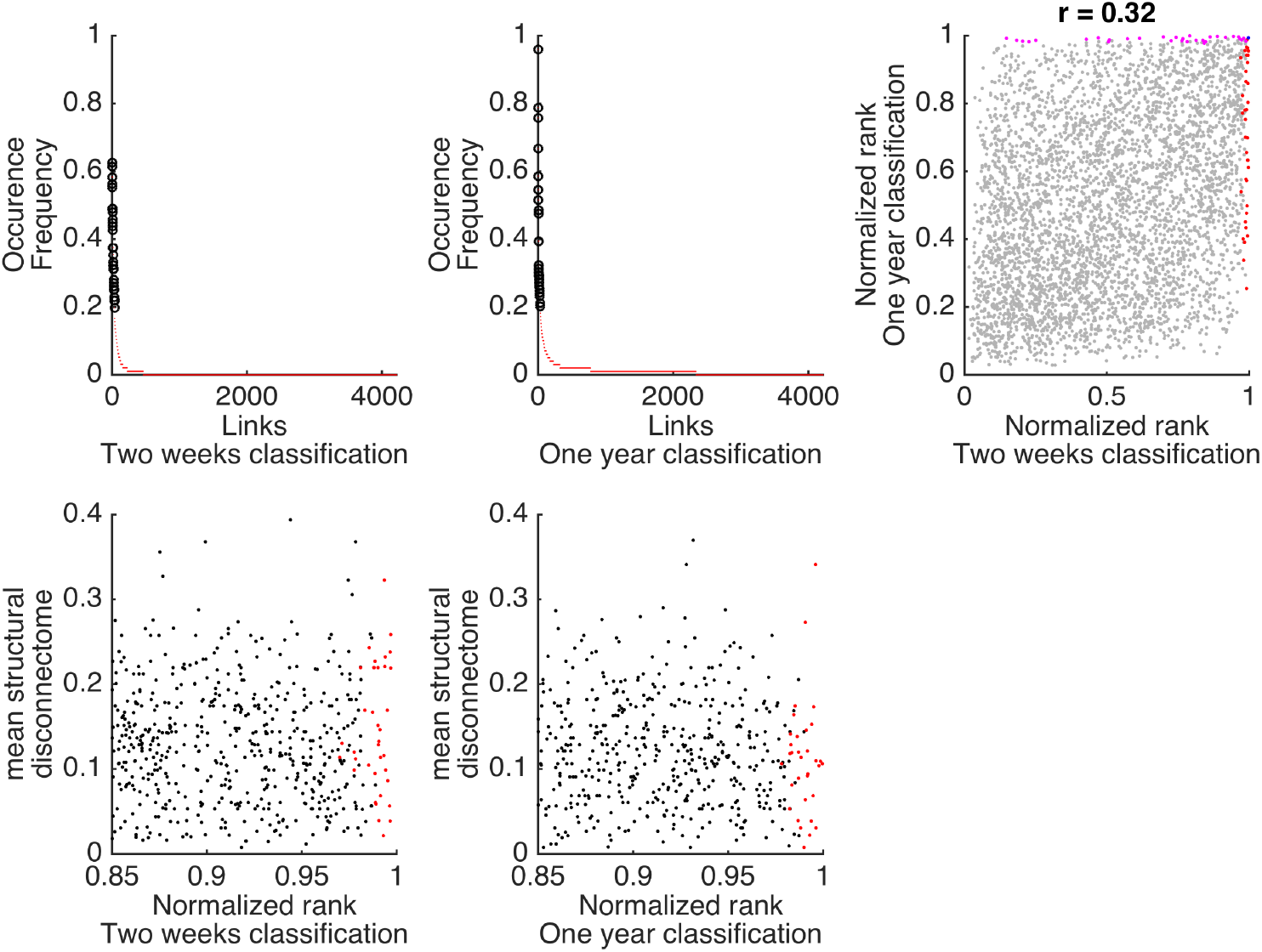
A, B: Frequency of occurrence in the predictive set for each link in the HCP-SCT across all train-test splits for classification of patients according to the number of behavioral deficits at the two weeks (A) and the one year (B) time points. Only a small fraction – 11% and 9.7%, respectively, are predictive in case of at least one split for the two weeks and the one year time points respectively. Black circles indicate the links that feature in the final selection as they occur in at least 20% of all train-test splits. C: Comparison of normalized ranking averaged across all train-test splits for all HCP-SCT links in the classification of patients at the two weeks and one year time points. Predictive links for individual classifications mentioned along x and y axes are displayed in red and magenta respectively while the common ones are shown in blue. D, E: Comparison of normalized ranking, averaged across all train-test splits, for highest ranked links in the classification of patients at the two weeks (D) and the one year (E) time points with structural disconnection for each link, averaged across patients. Predictive links in each case are shown in red. In none of the cases, a significant correlation between the ranking and average structural disconnection is found, whether all HCP-SCT links or only the predictive ones are considered. Here two weeks EC values for all links have been used for classification at both time points as they showed significantly higher accuracy than the chance level (Figure 5).

**Figure S9.**
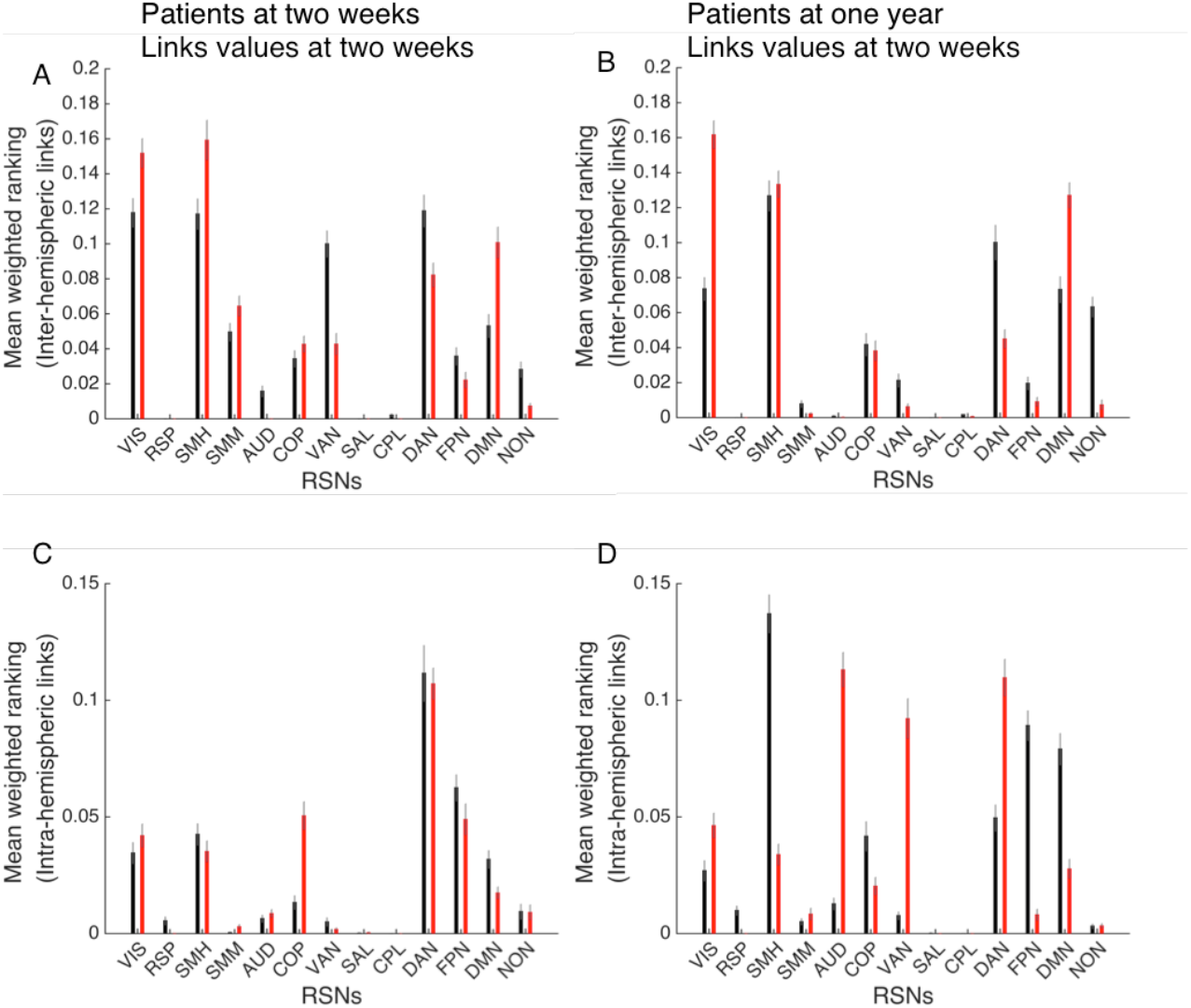
Mean (+/- SEM) weighted ranking of efferent (black) and afferent (red) EC predictive links per RSN for classification of patients according to the number of deficits at the two weeks (A, C) and one year (B, D) time points. Top panels consider only the inter-hemispheric links while the bottom panels consider only intra-hemispherical links. In all cases, only EC values at two weeks are considered as they showed significantly higher accuracy than the chance level (Figure 5).

## Notes

### Competing Interest Statement

The authors have declared no competing interest.

### Funding Statement

Funding was provided by NIH grant R01 NS095741 to M.C.; M.G. was supported by Horizon 2020 Framework Programme, Award ID: Human Brain Project SGA2 No. 785907.

### Author Declarations

Study procedures were performed in accordance with the Declaration of Helsinki ethical principles and approved by the Institutional Review Board at Washington University in St. Louis. The complete data collection protocol is described in full detail in a previous publication (Corbetta et al., Neuron, 2015).

